# Whole Exome Sequencing Reveals *FCGBP* Variant Associated with Spontaneous Intraabdominal Hemorrhage in Severe Acute Pancreatitis

**DOI:** 10.1101/2024.08.06.24311443

**Authors:** Qiu-Yi Tang, Yue-Peng Hu, Qi Yang, Jing Zhou, Jing-Zhu Zhang, Jie Yang, Haibin Hao, Gang Li, Bai-Qiang Li, Lu Ke, Zhi-Hui Tong, Yu-Xiu Liu, Evan Yi-Wen Yu, Wei-Qin Li

## Abstract

This study sought to identify genetic cause of spontaneous intraabdominal hemorrhage (SIH) in severe acute pancreatitis (SAP) to develop more effective treatment for this life-threatening complication. A four-phase study was conducted, leveraging a large-scale acute pancreatitis (AP) patients (n=600); the first phase involved whole-exome sequencing analyses, and identified specific exonic variant located in *FCGBP* (i.e., rs1326680184) that was consistently associated with SIH; the second phase performed serum ELISA tests, and revealed that *FCGBP* variant altered FCGBP level and further led to predisposition of SIH; the third phase conducted an i) *in-vivo* experiment with a *Fcgbp*-knockdown mouse model, and demonstrated lower expression of *Fcgbp* led to more severe AP morphology and higher risk of hemorrhage; ii) *in-vitro* experiment with *FCGBP*-knockdown human vascular fibroblasts demonstrated that down-regulated *FCGBP* expression could destabilize the vascular wall, and lead to vascular injury in SAP; the fourth phase compared *FCGBP* variant carriers to non-carriers with clinical characteristics, and found *FCGBP* variant associated with higher risks of poor complications and AP prognosis and enhanced the diagnostic capability as an indicator. These findings provide important insights into the underlying mechanism of SIH in SAP, and facilitate therapeutic development for AP prognosis and critical care in an early phase.

**Highlights:**

- Genetic mutation in *FCGBP* presents a strong association with predisposition of spontaneous intraabdominal hemorrhage, and provide a novel insight in increasing the severity of acute pancreatitis when knockdown the expression of Fcgbp.
- The incorporation of *FCGBP* mutation as an indicator enhances the ability of clinical assessment with respect to complications and mortality of acute pancreatitis in an early phase before manifestation.
- Our findings highlight the gene *FCGBP* as a probable pathogenic cause of spontaneous intraabdominal hemorrhage in severe acute pancreatitis patients, which enable a development of effective targeted therapies in improving the prognosis and critical care of severe acute pancreatitis.

## Introduction

Acute pancreatitis (AP), an acute inflammatory condition of pancreas, has become one of the leading causes of acute abdominal diseases. The annual incidence of AP ranges from 13 to 45 per 100,000 people, with approximately 20.0% progressed to severe outcomes and even death.^1,2^ The mortality of AP, as pooled from population-based cohort studies, is 1.16 per 100,000 general population per year.^3^ Nevertheless, the severe form of AP (SAP), which can lead to complications such as infected pancreatic necrosis (IPN) and acute respiratory distress syndrom (ARDS), has been reported to be associated with a high mortality rate over 30.0%.^4^ Unfortunately, there are currently no therapeutic agents available that can alter the course of AP, and the mechanisms of those complications remain largely unknown.

As one of the life-threatening complications, intraabdominal hemorrhage occurs during the course of AP, particularly SAP, and results in an elevated risk of abdominal infection, multiple organ dysfunction and other fatal consequences. While multifactorial characteristics, e.g., progressive inflammation, portal hypertension, necrosis and surgical necrosectomy, have been reported to be related to intraabdominal hemorrhage,^5^ the evidence remains controversial. The causes and pathogenesis of intraabdominal hemorrhage, particularly of spontaneous intraabdominal hemorrhage (SIH), have not been well understood. SIH is a critical form of intraabdominal hemorrhage that occurs in the course of AP without prior surgical intervention to the pancreas, making it difficult to be recognized and managed in the early phase of AP.

According to Chen *et al.*,^6^ AP patients complicated with SIH had a worse prognosis, with a higher mortality (54.2%) than those without SIH (20.8%). Therefore, identifying high-risk AP patients before this condition manifest is crucial. To our best knowledge, merely studies have investigated SIH in AP patients, while the risk factors and underlying mechanisms remain largely unknown. Given that previous studies have raised the suggestions for genetic variants as causative factors for several complications of AP,^7–11^ the role of genes, therefore, may be essential in the development of SIH.

In the past decade, next-generation sequencing techniques, including whole exome sequencing (WES), has been increasingly used to investigate genetic causes in complex diseases, particularly for agnogenic phenotypes.^12^ WES offers significant value in addressing unexplained presumed disorders by providing an opportunity to investigate potentially actionable variants and maximize clinical and biological exploration. This approach can guide clinical decision-making and provide valuable insights for selected experimental follow-up. Here we sought to use the largest WES study to date of AP to investigate the disease-specific influence of exonic variants on SIH to unveil potential cause and mechanism involved in SIH, thereby facilitating a more effective strategy for AP prognosis and critical care.

## Methods

### Participants enrollment

All the patients related to AP and admitted to Jinling Hospital, a tertiary level referral center in Nanjing, China, from 1^st^ January 2015 to 31^st^ December 2020, were retrospectively reviewed for eligibility in this study. All participants were informed about the study’s purpose during the admission interview and provided with consent prior to enrollment. The study design and conduct complied with all relevant regulations regarding the use of human participants and was in accordance with the criteria set by the Declaration of Helsinki. The study protocol was approved by the Ethics Committee of Jinling Hospital, Nanjing, China (ref. No. 2021NZKY-042-01). To definitude SAP complicated with SIH (i.e., SIH-SAP), all the participants were assessed according to stringent inclusion and exclusion criteria For inclusion criteria; i) diagnosed as AP according to the revised Atlanta criteria (RAC),^13^ ii) attributed as severe or critical AP according to determinants-based classification (DBC) guideline,^14^ iii) presence of intraabdominal hemorrhage on contrast-enhancing computed tomography without prior invasive intervention. For exclusion criteria; i) participants who had a history of hemophilia, idiopathic thrombocytopenic purpura, abdominal aorta aneurysm, or other hemorrhage associated disorders, ii) who received anticoagulation therapies within a week before admission.

The primary control group of SAP patients without SIH (i.e., non-SIH-SAP) was selected using a 1:1 ratio propensity score matching (PSM) based on the severity of AP. The severity was quantified based on Acute Physiology and Chronic Health Evaluation Ⅱ (APACHE Ⅱ) score and classified as mild, moderate, severe or critical according to DBC guideline.^14^ Additionally, to further validate the generalizability and stability of identified signals in SIH-SAP patients, a secondary control group was established consisting of AP patients without SIH (i.e., non-SIH-AP). Since 4 patients rejected to sign the informed consent form for using their blood to conduct WES, a total of 45 SIH-SAP patients, 45 non-SIH-SAP patients, and 510 non-SIH-AP patients were included in the current study. Clinical and anthropometric information were obtained from medical interviews, physical examinations and laboratory assessments at admission.

### Whole exome sequencing

Peripheral blood samples were collected within one day after admission and stored at -80°C. Genomic DNA of all participants was extracted from stored peripheral blood using TIANGEN kit (Beijing, China), following the manufacturer’s protocols. Whole-exome libraries were generated according to the manufacturer’s protocols, with enrichment of exon-coding regions using Agilent’s V6 capture reagent (Agilent Technologies, Santa Clara, CA). Paired-end (2×150 base pairs) sequencing was performed on Illumina NovaSeq 6000 System (Illumina, San Diego, CA) (please see details in Supplementary methods).

#### Variant-based analysis

After quality control and variant screening (as described in supplementary methods), a total of 1,322 candidate variants, including the uncommon variants (MAF <0.01) and variants without population alle frequency annotation, were selected for further analysis due to their potential pathogenic implications. Here, the candidate variants with different allele frequency between the case group (45 SIH-SAP patients) and the primary control group (40 non-SIH-SAP patients) were identified as probable pathogenic variants at *p*__fdr_<0.05 (*Fisher’s* exact method of R package “*stats*” and FDR correction with Benjamini-Hochberg method). In addition, the secondary control group consisting of 510 non-SIH-AP patients, was used to validate the identified exonic variants, in which the *Fisher’s* exact method was performed to assess the distribution of allele frequency compared to the case group of 45 SIH-SAP patients at *p*__fdr_<0.05. As a sensitivity analysis, all the 1,037,733 variants that passed quality control were also performed variant-based analysis according to the process mentioned above.

#### Targeted genome-wide association analysis

Based on the identified signals, a chromosome-19-specific analysis was performed to minimize the effects of genome-wide multiple testing. The high-quality variants from the case (i.e., SIH-SAP) group and control (i.e., non-SIH-SAP) group underwent stringent quality control measures and were subsequently merged. The variants from both case and control groups were filtered using PLINK (version 1.9.0)^15^ to ensure reliability. A total of 43,486 variants located in chromosome 19 were deemed eligible. Samples were excluded if they failed genotyping in more than 40% of variants. The variants were excluded if they met any of the following criteria: i) call rate <60%; ii) MAF <0.01; iii) significantly deviated from Hardy-Weinberg equilibrium (HWE) with *p*<1×10^-8^.^16^ The significance threshold was set at an exome-wide level of *p*<5×10^-6^ (1×10^-8^/1%, as exome accounts for approximately 1% of human genome). PLINK (version 1.9.0) was used to perform this chromosome-19-specific targeted genome-wide association analysis, controlling for age (years) and gender (male or female) as covariates. Also, an exome-wide association analysis was performed with variants detected across all chromosomes to account for any unexpected signals, applying the same statistical models and criteria used in the chromosome-19-specific analysis.

#### Gene-based analysis

Variants with MAF <0.01 or lacking allele frequency information in genome reference databases (i.e., Exome Aggregation Consortium and 1000 Genomes) and predicted to be deleterious, were subjected to gene annotation. A sequence kernel association test (SKAT),^17,18^ a gene-based analysis, was employed to explore the associations between genes containing at least two exonic variants, such as *FCGBP*, and development of SIH in all the included participants. Since not all the variants defined as “uncommon” have a MAF <1% in our SIH-SAP patient group, a different approach was applied, which is to define the variants with 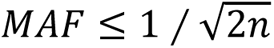 as “uncommon” whereas variants with 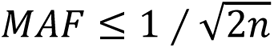 as “common” (n is the total sample size).^17^ The combined effects of uncommon and common exome variants on SIH were assessed by using R package “*SKAT*”.

### Experiments

All animal experiment procedures followed Chinese Animal Welfare Act, the Guidance for Animal Experimentation of Southeast University. Efforts were made to minimize the number of animals used and reduce their suffering (ref. No. 2022DZGKJDWLS-

0059). Male ICR mice aged 8 weeks were obtained from Ziyuan Laboratory Animal Technology Co., Ltd. (Hangzhou, China). Animals were housed and maintained in specific pathogen-free (SPF) facilities with a 12-hour light-dark cycle and free access to standard food and water.

#### Knock-down of Fcgbp

*Fcgbp* was silenced using an adeno-associated virus (AAV) vector delivery system. An AAV vector harboring a short hairpin RNA (shRNA) targeting *Fcgbp* (Refseq NM_001122603.1) (AAV-RNAi) was provided by Gene Chem Co., Shanghai, China. For *Fcgbp*-target AAV-RNAi packaging, the designed shRNA primers (Supplementary Table 1) were inserted into the vector. The titer of AAV was 3.36×10^13^ viral genome copies per ml, and a total of 5.00×10^11^ viral genome copy was infused through intravenous injection per mouse.

#### In-vivo acute pancreatitis mice model

The SAP mouse model was induced by intraperitoneal injection of caerulein (50 μg/kg for 7 times with one-hour intervals) followed by lipopolysaccharide (administered one hour after the final caerulein injection at 5mg/kg, one time).^19^ The pathological examination of pancreas in SAP mice and normal mice are presented in Supplementary Fig. 1. The blood assays for TNFα, IL-6, MCP1 are provided in Supplementary Fig. 2. The *Fcgbp-KD* mice, together with the mice injected with either the AAV vector or PBS, were induced as SAP on the 30^th^ day since the day of AAV or PBS injection (n=10 for each group). At 24h post-SAP induction, the mice were anesthetized, and tissues of lung, pancreas, colon, and vessel were harvested before the mice were sacrificed with minimal suffering.

#### Immunohistochemistry

Immunohistochemistry (IHC) was performed to in terms of Fc Fragment of Immunoglobulin G Binding Protein (Fcgbp) in lung, colon and vessel tissues that were obtained from the sacrificed mice. Briefly, 5-μm-thick cryosections of quick-frozen lung, colon and vessel tissues were fixed in acetone, quenched with 3% H_2_O_2_, and blocked with goat serum. After washes with PBS, the sections were treated with anti-*Fcgbp* primary antibody (bs-13168R, Bioss, Beijing, China) for 2 hours. Then, the sections were incubated with horseradish-peroxidase-conjugated secondary antibody for an hour. Color development was achieved using 3,3’-diaminobenzidine tetra hydrochlorides as peroxidase substrate. The proportion of Fcgbp with positive response was quantified using ImageJ. The difference of positive areas in three groups (i.e., control, AAV-RNAi and AAV vector groups) were compared using one-way ANOVA with post-hoc testing to evaluate the significance of difference among three groups.

#### Immunofluorescence

Immunofluorescence was conducted in terms of Fcgbp, Cd31, α-Sma, and Vegf in vessel tissues that were obtained from the sacrificed mice. The procedure followed the same initial steps as immunohistochemistry up to the secondary antibody incubation. After washing three times with PBS, the tissue was reacted with fluorescent dye.

#### Morphological examination

Fresh pancreatic and pulmonary tissue samples were fixed with 4% paraformaldehyde overnight. After fixation, the samples were washed with running water for 2 hours, dehydrated with a gradient of ethanol, embedded in paraffin, and then cut into 5-μm-thick sections. The sections were dewaxed in xylene, rehydrated through downgrade ethanol solutions (100%, 95%, 80%, 70%), and stained with hematoxylin and eosin. Morphological examination, including edema area and grade, acinar cell necrosis, inflammatory reaction, adipose necrosis, and hemorrhage, was assessed to determine the severity of AP for each group in mice (n=10 per group) by two independent and blinded investigators using previously reported morphometry methods.^20^ The differences of severity scores among the three groups were evaluated using one-way ANOVA with post-hoc tests.

#### Cell lines and cultures

Cell line of human vascular fibroblast was obtained from Wuhan Sunncell Biotech Co., Ltd. (SNP-H362, Hubei, China) and cultured in a specific complete culture medium provided by Wuhan Sunncell Biotech Co., Ltd. (SNPM-H362, Hubei, China). The cells were maintained in a 5% CO_2_ humidified incubator at 37°C in media supplemented with 10% fetal bovine serum (Gibco, Grand Island, NY, USA) and 1% penicillin and streptomycin (Gibco, Grand Island, NY, USA).

#### Construction of knockdown cell

Cells designated for knockdown construction were thoroughly digested and plated in 6-well plate at a density of 70–80%. *FCGBP*-targeting siRNAs were then introduced and incubated. The efficiency of the knockdown was assessed by qRT-PCR and western blot analysis.

#### RNA-Seq and bioinformatics analysis

Total cellular RNA was extracted from both the knockdown fibroblasts and the wild-type controls, with residual DNA removed using DNase I (Roche Diagnostics). RNA-seq libraries were prepared using the TruSeq RNA Library Prep Kit and sequenced on the Illumina NextSeq 500 platform. The resulting data were processed to compute transcript FPKMs. Pathway analysis was performed using the GO (http://geneontology.org/) and KEGG (https://www.genome.jp/kegg/) databases.

### Investigation of identified variant to clinical characteristics

AP patients in case group, primary control group, and secondary control group were pooled (n=600) irrespective of disease severity and presence of SIH. These patients were then re-categorized according to genetic status of the identified variant as carriers or non-carriers. To link the identified variant(s) to clinical characteristics, analyses were conducted comparing variant carriers to non-carriers. This involved comparing the distribution of clinical characteristics and testing their associations with the identified variant using a standard logistic regression adjusted for age (years, continuous), gender (male or female), BMI (Kg/m^2^, continuous) and etiology of AP (biliary, hypertriglyceridemia, and others). Blood test parameters were measured during the first blood draw upon admission to the hospital. The receiver operating characteristic curve (ROC) (R package “*pROC*”),^21^ and DeLong’s test^22^ for comparison of area under the ROC curve (AUROC) were employed to assess whether the identified variant increase the diagnostic ability for a series of clinical outcomes, including SIH, in addition to demographic and clinical factors.

#### Statistical analysis

Descriptive statistics are presented as median (interquartile range) for continuous variables and frequency (percentage, %) for categorical variables. Demographic and clinical characteristics between different groups of participants were compared by Wilcoxon rank-sum test for continuous variables, and *Fisher’s* exact test for categorical variables. All the statistical analyses were performed using R software 4.1.0 (R Core Team. R Foundation for Statistical Computing. Vienna, Austria. 2021. https://www.R-project.org).

#### Statistical power and multiple hypothesis testing

To minimize the occurrence of false discoveries, the current study utilized a series of measures as: i) genetic variants with relatively low reliability were eliminated based on quality indicators including missing rate, quality score, and VQSR; ii) a filtration strategy was applied to restrict the number of variants tested, excluding common and synonymous variants; iii) a false discovery rate correction was used to account for multiple testing in the comparison of allele frequency distribution among the included candidate variants; iv) to ensure the reliability and reproducibility, the aforementioned analyses were replicated with a secondary control group of non-SIH-AP patients to verify the consistency of the identified genetic signals.

## Results

### Demographic and clinical characteristics of included participants

A detailed description of the study design, workflow, and data processing is displayed in Fig. 1. Among patients admitted with AP to a tertiary medical center (Jinling hospital, Nanjing, China), a total of 49 patients were determined in a severe or critical scenario of AP and diagnosed with SIH (SIH-SAP). No pedigree relatedness was observed according to the lineage data gained from all included participants. The primary control group of SAP patients without SIH (non-SIH-SAP) was selected using a 1:1 ratio propensity score matching (PSM) based on severity of AP, which were quantified using the Acute Physiology and Chronic Health Evaluation Ⅱ (APACHE Ⅱ) score and classified as mild, moderate, severe or critical according to determinants-based classification (DBC) guideline.^14^ As showed in Table 1, no difference between the case group (i.e., SIH-SAP patients) and the primary control group (i.e., non-SIH-SAP patients) in terms of the median of APACHE Ⅱ score (15.00 vs. 15.00), incidence rates of acute kidney injury (80.0% vs. 68.8%) and acute respiratory distress syndrome (82.2% vs. 80.0%) were observed. The median (interquartile range) age at admission was 46 (33–56) years for the case group, and 38 (30–44) years for the primary control group (*p*=0.007). Females comprised 17.8% of the case group and 46.7% of the primary control group (*p*=0.006). The mortality rate of SIH-SAP patients (35.6%) is extensively higher than that of non-SIH-SAP patients (22.2%), while no significant difference was observed. In addition, results of coagulation functional test showed a higher thrombin time (median seconds; interquartile range) in the case group (17.00; 15.90–18.30) than the primary control group (15.70; 14.80–17.40; *p*=0.027), and a lower serum fibrinogen (median g/L, interquartile range) in the case group (4.09; 2.76–4.79) than the primary control group (4.56; 3.84–5.90; *p*=0.022). The results of thrombin time indicated SIH-SAP patients tended to have a fibrinogen deficiency or dysfunctional fibrinogen; similarly, the lower serum fibrinogen, a precursor of fibrin that is crucial for formation of blood clot in tissue or vascular injury, suggested a lightly poorer coagulation function of SIH-SAP patients.

**Fig. 1.**
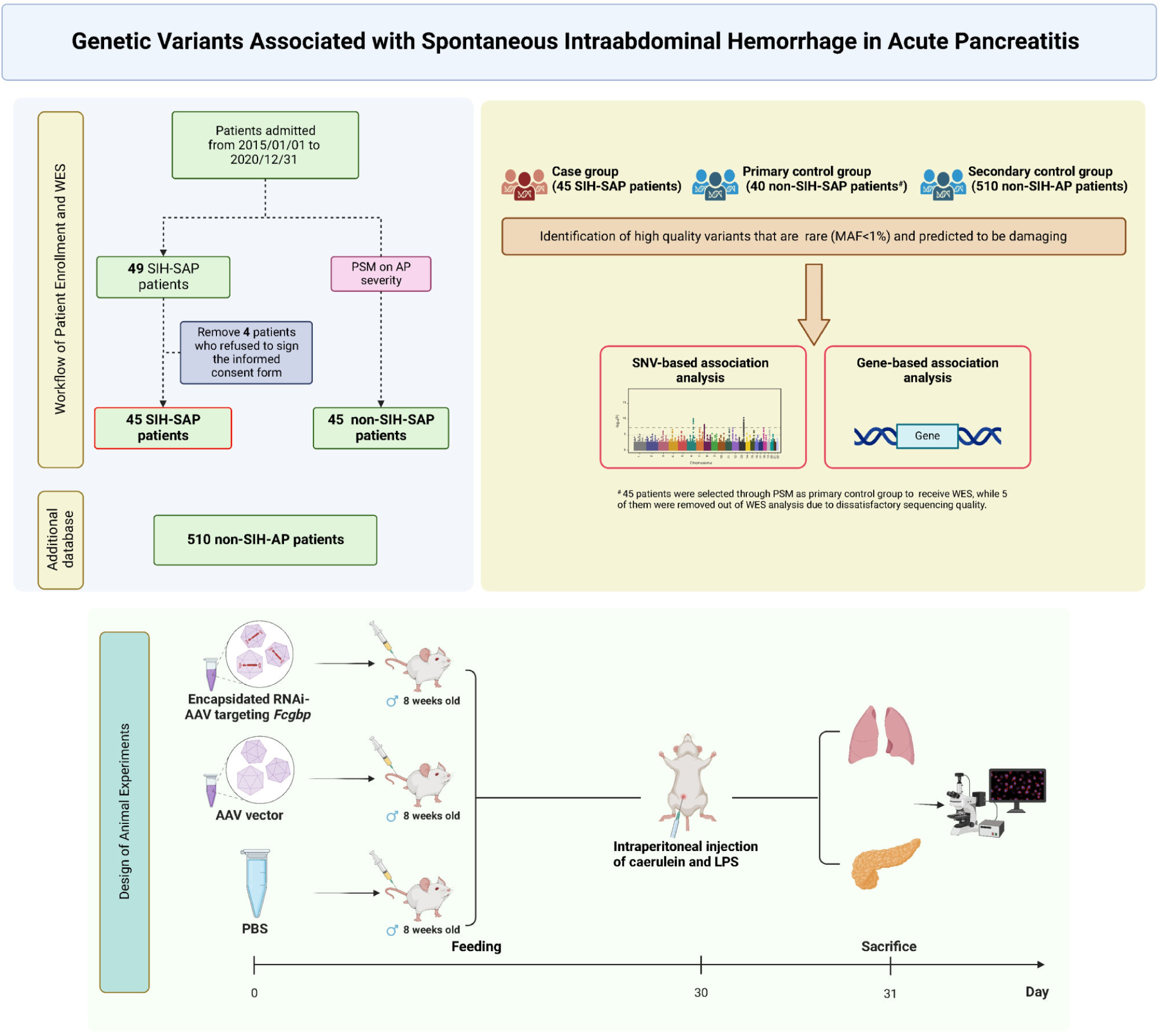
The schematic design and workflow of participants enrollment, bioinformatics analysis, and animal experiments. In total, 45 SIH-SAP cases, 45 primary non-SIH-SAP controls and 510 secondary non-SIH-AP controls were included in the current study. A series of analyses were performed to identify the exome variants related to SIH development. Animal experiments were used to verify the effects of identified gene on causing SIH in SAP model. Abbreviations: MAF, minor allele frequency; RNAi-AAV, adeno-associated virus packaged with small hairpin RNA; LPS, lipopolysaccharides; PBS, phosphate buffer saline; SIH, spontaneous intraabdominal hemorrhage; AP, acute pancreatitis patients; SAP, severe acute pancreatitis.

**Table 1.**
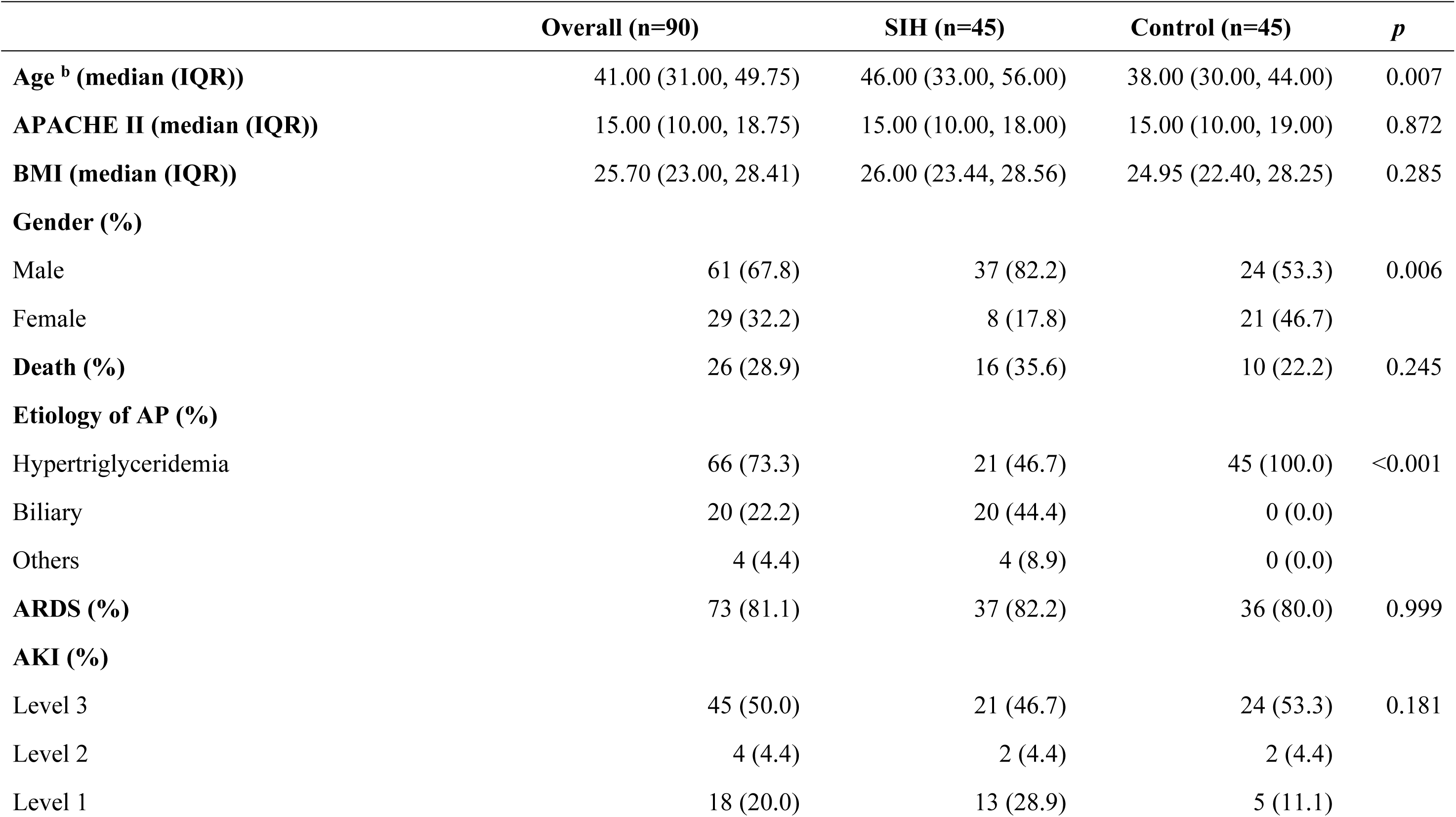
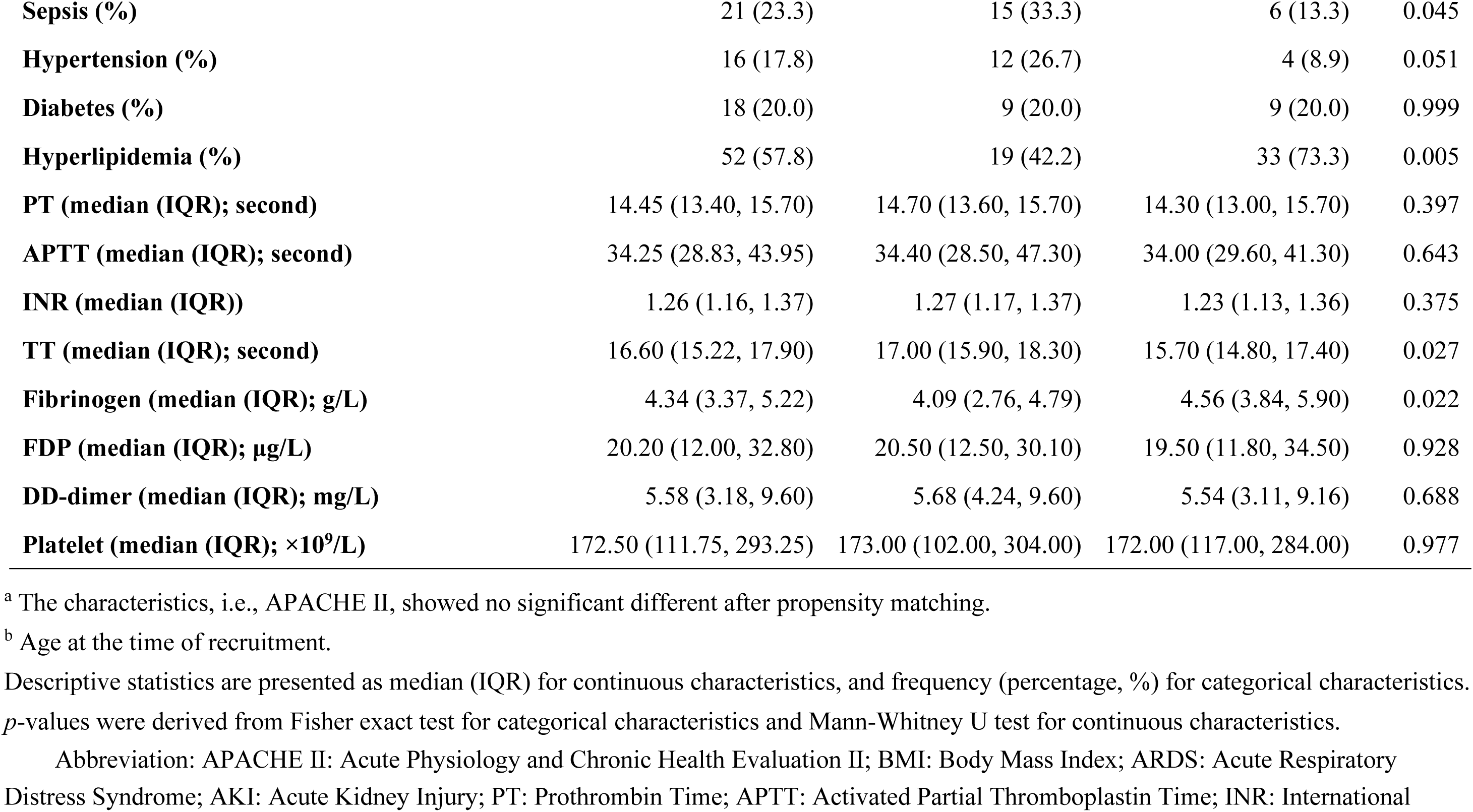
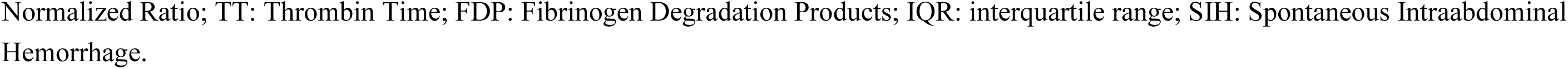
Demographical and clinical characteristics of participants ^a^.

|  | **Overall (n=90)** | **SIH (n=45)** | **Control (n=45)** | ***p*** |
| --- | --- | --- | --- | --- |
| **Age <sup>b</sup> (median (IQR))** | 41.00 (31.00, 49.75) | 46.00 (33.00, 56.00) | 38.00 (30.00, 44.00) | 0.007 |
| **APACHE II (median (IQR))** | 15.00 (10.00, 18.75) | 15.00 (10.00, 18.00) | 15.00 (10.00, 19.00) | 0.872 |
| **BMI (median (IQR))** | 25.70 (23.00, 28.41) | 26.00 (23.44, 28.56) | 24.95 (22.40, 28.25) | 0.285 |
| **Gender (%)** |  |  |  |  |
| Male | 61 (67.8) | 37 (82.2) | 24 (53.3) | 0.006 |
| Female | 29 (32.2) | 8 (17.8) | 21 (46.7) |  |
| **Death (%)** | 26 (28.9) | 16 (35.6) | 10 (22.2) | 0.245 |
| **Etiology of AP (%)** |  |  |  |  |
| Hypertriglyceridemia | 66 (73.3) | 21 (46.7) | 45 (100.0) | <0.001 |
| Biliary | 20 (22.2) | 20 (44.4) | 0 (0.0) |  |
| Others | 4 (4.4) | 4 (8.9) | 0 (0.0) |  |
| **ARDS (%)** | 73 (81.1) | 37 (82.2) | 36 (80.0) | 0.999 |
| **AKI (%)** |  |  |  |  |
| Level 3 | 45 (50.0) | 21 (46.7) | 24 (53.3) | 0.181 |
| Level 2 | 4 (4.4) | 2 (4.4) | 2 (4.4) |  |
| Level 1 | 18 (20.0) | 13 (28.9) | 5 (11.1) |  |
| **Sepsis (%)** | 21 (23.3) | 15 (33.3) | 6 (13.3) | 0.045 |
| **Hypertension (%)** | 16 (17.8) | 12 (26.7) | 4 (8.9) | 0.051 |
| **Diabetes (%)** | 18 (20.0) | 9 (20.0) | 9 (20.0) | 0.999 |
| **Hyperlipidemia (%)** | 52 (57.8) | 19 (42.2) | 33 (73.3) | 0.005 |
| **PT (median (IQR); second)** | 14.45 (13.40, 15.70) | 14.70 (13.60, 15.70) | 14.30 (13.00, 15.70) | 0.397 |
| **APTT (median (IQR); second)** | 34.25 (28.83, 43.95) | 34.40 (28.50, 47.30) | 34.00 (29.60, 41.30) | 0.643 |
| **INR (median (IQR))** | 1.26 (1.16, 1.37) | 1.27 (1.17, 1.37) | 1.23 (1.13, 1.36) | 0.375 |
| **TT (median (IQR); second)** | 16.60 (15.22, 17.90) | 17.00 (15.90, 18.30) | 15.70 (14.80, 17.40) | 0.027 |
| **Fibrinogen (median (IQR); g/L)** | 4.34 (3.37, 5.22) | 4.09 (2.76, 4.79) | 4.56 (3.84, 5.90) | 0.022 |
| **FDP (median (IQR); µg/L)** | 20.20 (12.00, 32.80) | 20.50 (12.50, 30.10) | 19.50 (11.80, 34.50) | 0.928 |
| **DD-dimer (median (IQR); mg/L)** | 5.58 (3.18, 9.60) | 5.68 (4.24, 9.60) | 5.54 (3.11, 9.16) | 0.688 |
| **Platelet (median (IQR); ×10<sup>9</sup>/L)** | 172.50 (111.75, 293.25) | 173.00 (102.00, 304.00) | 172.00 (117.00, 284.00) | 0.977 |

<sup>a</sup> The characteristics, i.e., APACHE II, showed no significant different after propensity matching.
<sup>b</sup> Age at the time of recruitment.
Descriptive statistics are presented as median (IQR) for continuous characteristics, and frequency (percentage, %) for categorical characteristics. *p*-values were derived from Fisher exact test for categorical characteristics and Mann-Whitney U test for continuous characteristics.
Abbreviation: APACHE II: Acute Physiology and Chronic Health Evaluation II; BMI: Body Mass Index; ARDS: Acute Respiratory Distress Syndrome; AKI: Acute Kidney Injury; PT: Prothrombin Time; APTT: Activated Partial Thromboplastin Time; INR: International
Normalized Ratio; TT: Thrombin Time; FDP: Fibrinogen Degradation Products; IQR: interquartile range; SIH: Spontaneous Intraabdominal Hemorrhage.

### Whole exome sequencing and candidate variants selection

Except for the sequencing data of 5 patients in the primary control group that failed to meet the quality requirements and thereby were removed out of the WES analysis, all the rest of samples produced high-quality WES data with >80.0% of quality score ≥Q30 (99.9% of base call accuracy; Supplementary Table 2). On average, 41 million reads per sample (range 35–51 million) were generated, each with an average read length of 150bp. Over 99.9% of reads for each sample were on target and 8,334,678 variants were obtained. Nevertheless, a total of 1,037,733 variants (12.5%) remained after quality control. Those variants were then annotated and screened for variants of interest (i.e., minor allele frequency (MAF) <1% and nonsynonymous), which maintained 1,322 candidate variants for further analyses (Fig. 2A). Principal components analysis revealed a homogenous population structure between the SIH-SAP patients and non-SIH-SAP patients (*p*=0.633) (Fig. 2B).

**Fig. 2.**
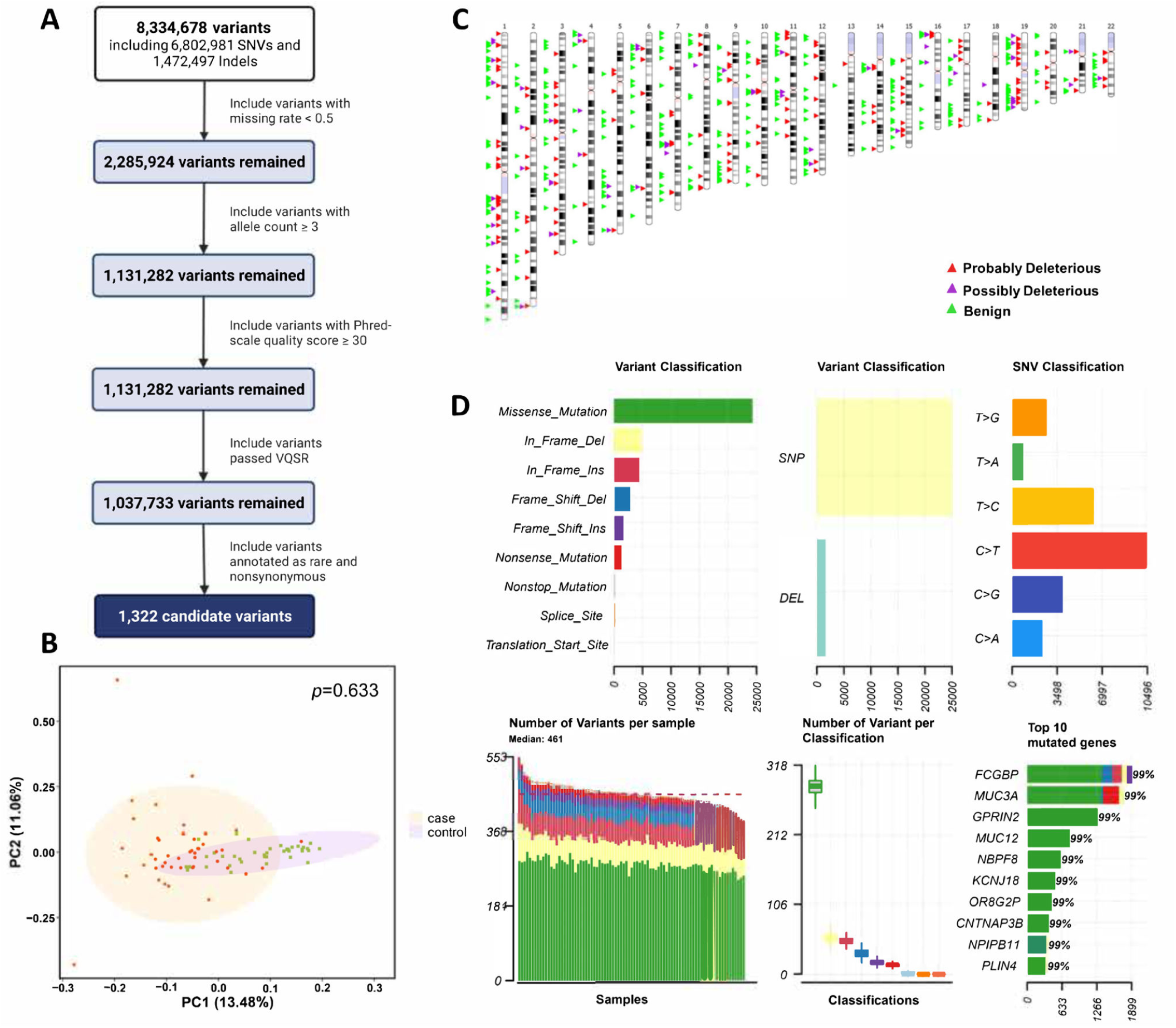
Overview of candidate variants selection and characteristics profiling. (**A**) flow-chart of inclusion and exclusion of genetic variants, 1,322 exonic variants were finally selected as candidate variants. (**B**) genetic principal component analysis of the included participants. (**C**) the distribution of predicted deleterious or benign candidate variants based on karyotype according to Sorting Intolerant from Tolerant (SIFT) algorithm. The red, purple and green triangles denote probably deleterious sites, possibly deleterious sites and benign sites, respectively. (**D**) summary characteristics of candidate exonic variants. Abbreviations: VQSR, variant quality score recalibration; SNV, single nucleotide variants; SNP, single nucleotide polymorphism; DEL, deletion; SIFT, Sorting Intolerant from Tolerant.

Nearly 22.3% (i.e., 295 variants) were predicted to be “probably deleterious” or “possibly deleterious” based on Sorting Intolerant from Tolerant algorithm (SIFT) (Fig. 2C), and 16.6% (i.e., 220 variants) were predicted to be “probably damaging” or “possibly damaging” based on Polymorphism Phenotyping (PolyPhen). The average number of candidate variants per sample was 461, with *FCGBP* having the highest number of variants among the set of variants analyzed (Fig. 2D). Comprehensive annotation of the candidate variants (Supplementary Table 3-12), the majority of 1,322 candidate variants were classified as missense (63.8%), followed by in-frame deletion/insertion (20.0%) and frame-shift deletion/insertion (13.9%). Many of these variants were associated with enhancer histone marks and epigenetic alteration. Specifically, 367 variants were observed to be located in the sites of the CpG island, indicating potential DNA methylation and resulting epigenetic alterations. To biologically understand the genes mapped to identified variants, gene enrichment pathway analyses using GO database identified 68 significant pathways (*p*<0.05), primarily related to metabolism of protein and extracellular matrix organization.

### FCGBP variant associated with SIH-SAP

The allele distribution between the case group (i.e., SIH-SAP) and the primary control group (i.e., non-SIH-SAP) was compared to identify the potential pathogenic variants with significant different alternative allele frequencies between the two patient groups (*p___*_fdr_<0.05). Of these candidate variants, rs1326680184, which located in *FCGBP* (Refseq NM_003890), was identified as being associated with SIH development in SAP patients. In contrast to non-SIH-SAP patients (11 out of 40; 27.5%), a significantly higher proportion of SIH-SAP patients were found to harbor the *FCGBP* variant (36 out of 45; 80.0%; Fig. 3B). A analysis was conducted using the secondary control group, consisting of 510 AP patients confirmed as non-SIH (i.e., 510 non-SIH-AP patients) from a prospectively-registered cohort of AP patients in our center. The finding was validated by comparing the carriers of rs1326680184 in SIH-SAP patients (36 out of 45; 80.0%) and non-SIH-AP patients (192 of 510; 37.6%) (*p*__fdr_<0.001). The identification of the specific *FCGBP* variant maintained consistent when stratified for severity of AP, which indicated a similar carriers for 184 severe or critical AP patients (38.9%) and 326 mild or moderate AP (36.8%, p>0.05), suggesting an extensively conservative effect of the *FCGBP* variant on SIH irrespective of AP severity. Moreover, no additional variants were found to be associated with SIH development in the sensitivity analysis, which included all 1,037,733 variants that passed quality control. To address multiple testing in a relatively small sample size, we employed a targeted GWAS method focusing on signals within chromosome 19, with respect to SIH-SAP patients vs. non-SIH-SAP patients. Again, the identified *FCGBP* variant (i.e., rs1326680184) was found to be consistently associated with the development of SIH (*p*__adj_=3.855×10^-12^; Fig. 3C and Supplementary Fig. 3). Moreover, we conducted a whole-exome wide association test by analyzing exome variants across all chromosomes, revealing multiple subtle signals with *p*__adj_<5×10^-6^ (Supplementary Fig. 4).

**Fig. 3.**
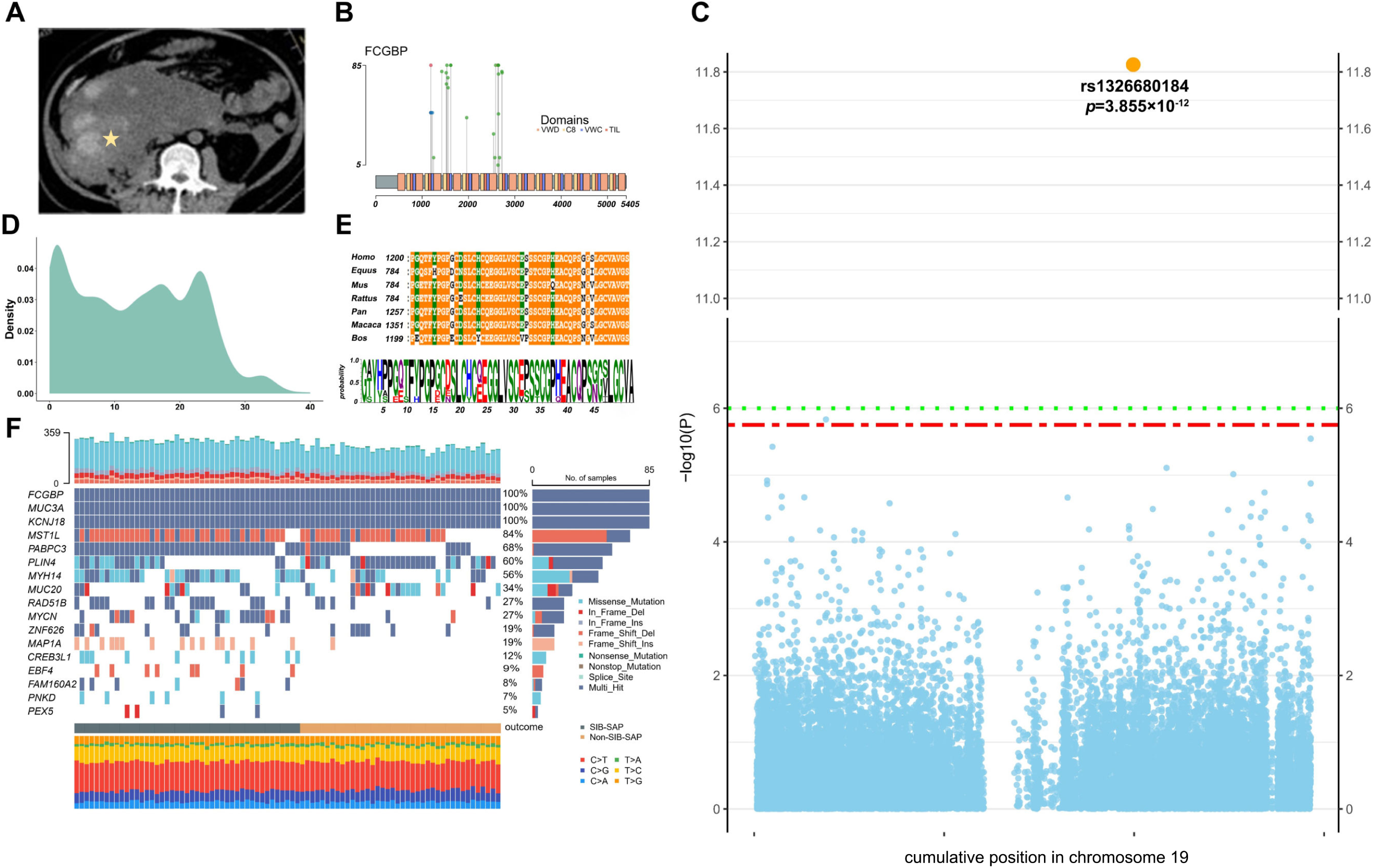
Bioinformatics analyses identify *FCGBP* and its variant to be associated with SIH development. (**A**) an example of computed tomography for SIH. The yellow pentagram indicates the focal lesion of SIH in an SAP patient. (**B**) distribution of *FCGBP* variants among SIH-SAP patients. A frameshift deletion is symbolized by a blue circle, whereas a green circle denotes an altered amino acid. (**C**) Manhattan plot of GWAS based on exome variants in chromosome 19. The x-axis is chromosomal position and the y-axis is –log_10_(*p* value of GWAS). The rs1326680184 was used and denoted to represent the six identified *FCGBP* variants. (**D**) distribution of CADD score of 1,322 candidate variants. (**E**) Protein sequence alignment of FCGBP in *Homo sapiens*, *Equus caballus*, *Mus musculus*, *Rattus norvegicus*, *Pan troglodytes*, *Macaca mulatta* and *Bos taurus*. The results revealed that the screened residues of *FCGBP* is highly conserved across multiple species and isoforms. In this alignment, residues that are absolutely conserved and highly conserved are highlighted in orange and green, respectively. (**F**) Gene-based variants of SAP patients. The matrix in the middle panel indicates each column representing per patient, and each row representing per gene. The bar plot on the top shows the number of variants in each individual and the bar plot on the right side shows the percentage of individuals containing a variant in each gene. The bottom plot shows the distribution of 6 types of variants per individual. Abbreviations: MAF, minor allele frequency; RNAi-AAV, adeno-associated virus packaged with small hairpin RNA; LPS, lipopolysaccharides; PBS, phosphate buffer saline; SIH, spontaneous intraabdominal hemorrhage; AP, acute pancreatitis patients; SAP, severe acute pancreatitis; FCGBP/Fcgbp, Fc Fragment of Immunoglobulin G Binding Protein; GWAS, genome-wide association study; CADD, combined annotation dependent depletion.

According to genomic annotation, the rs1326680184 is a frameshift deletion located at chromosome 19:39906058-39906060 within the exonic region of *FCGBP*. By using the combined annotation dependent depletion (CADD) score,^23^ rs1326680184 was predicted to be deleterious with a scaled C-scores of 20.7, indicating a likely negative effect (Fig. 3D). Additionally, conservative analysis suggests that the reginal residues of *FCGBP* are mostly conserved across species and isoforms (Fig. 3E).

Through SKAT analysis,^17,18^ 17 genes showed a suggestive association with SIH among SAP patients (*p<*0.01) (Supplementary Table 13). According to the rank of the number of patients carrying the candidate variants, *FCGBP* was again, identified as the top hit (*p*=1.310×10^-7^ in SKAT; Fig. 3F).

#### ELISA validation of FCGBP expression affected by FCGBP variant in AP patients

Enzyme-linked immunosorbent assay (ELISA) for FCGBP protein was conducted in peripheral blood of randomly selected AP patients, grouped by the presence (n=27) or absence (n=27) of the rs1326680184 variant. The results showed that AP patients with rs1326680184 had a lower level of FCGBP compared to those without rs1326680184 (mean of 2.86 ng/ml vs. 4.66 ng/ml, *p*=0.035, Fig. 4A). To verify the reduction of FCGBP levels resulting in predisposition of SIH, the comparison of FCGBP protein levels between SIH and non-SIH patients were conducted by grouping the presence (n=30) or absence (n=30) of SIH. The results suggest AP patients with SIH have a significantly lower level of FCGBP (*p<*0.001, Supplementary Fig. 5), which draw a consistency that *FCGBP* variant alter the FCGBP level, and lower FCGBP level leads to predisposition of SIH.

**Fig. 4.**
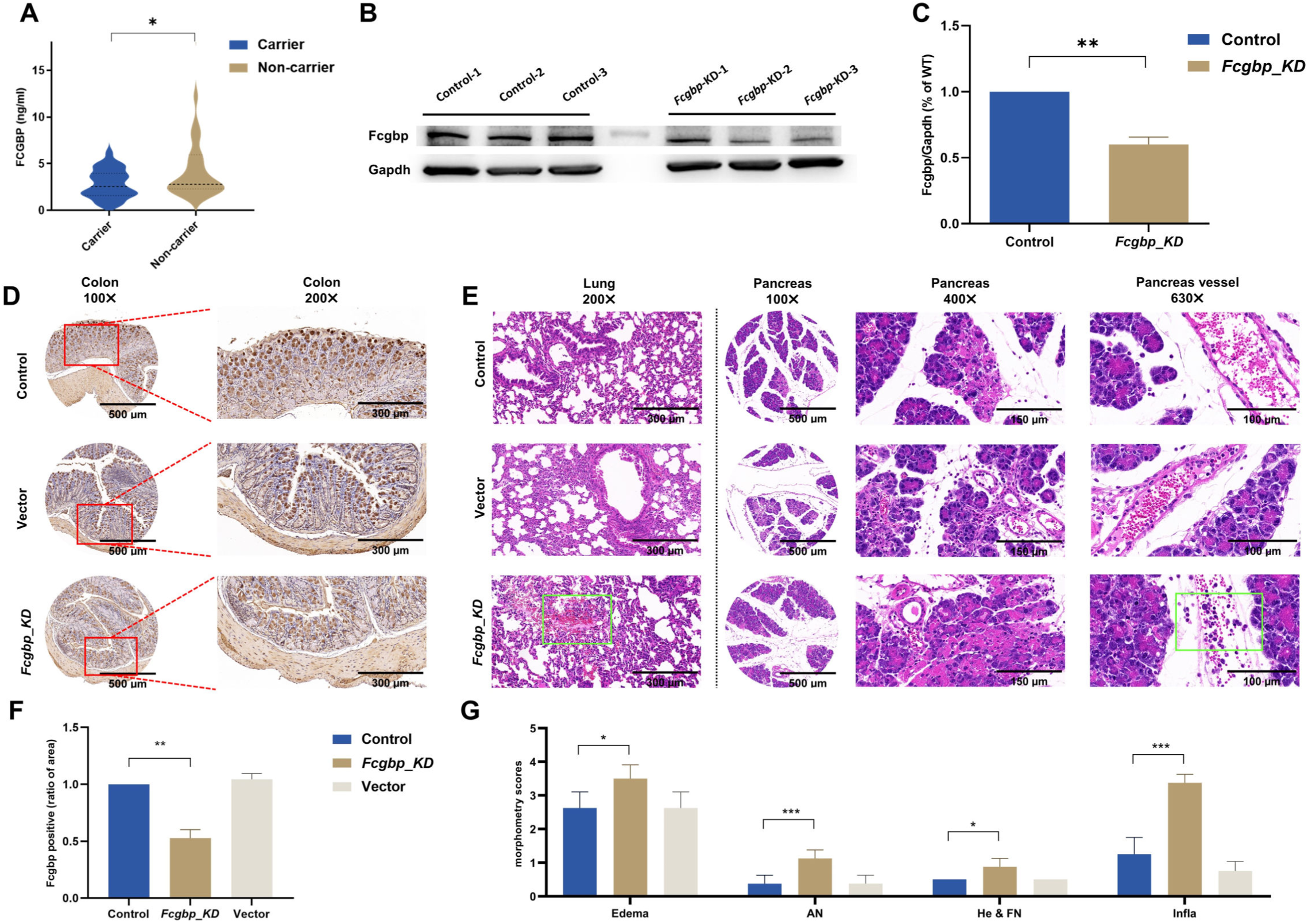
*FCGBP* variant (i.e., rs1326680184) relates to lower expression of *FCGBP* in patients and *Fcgbp-KD* exacerbates the severity and lung injury of AP in mice. (**A**) violin plot of protein FCGBP’s concentration in peripheral blood of AP patients, compared between rs1326680181 carriers and non-carriers (n=27 in each group), * indicates *p*<0.05 by *Student’s t* test with Welch’s correction. (**B**) western-blot upon protein Fcgbp from colon tissue of mice received *Fcgbp-KD* and normal healthy mice. (**C**) quantification of western-blot presents the ratio of Fcgbp/Gapdh (n=6 in each group), ** indicates *p*<0.01 by Man-Whitney test. (**D**) immunohistochemical analysis demonstrates the presence of Fcgbp in mice colonic tissue. Injection of AAV-RNAi targeting Fcgbp inhibits the expression of *Fcgbp*, compared with AAV empty vector and PBS as control (n=10 in each group). (**E**) quantification of *Fcgbp* expression. This was measured as stained area using ImageJ, taking control group as reference. ** indicates *p*<0.01 by one-way ANOVA with post-hoc tests. Error bars are shown as SEM. (**F**) morphological examination of lung and pancreas tissue (H&E staining). The results showed edema, acinar necrosis, hemorrhage and inflammation. Cellular edema and damage, infiltration of neutrophils is much severer in Fcgbp knock-down mice, and leakage of red blood cells were found in Fcgbp knock-down mice but not in the AAV empty vector injected mice and controls. (**G**) quantification of severity according to pathological scoring scale. * indicates *p*<0.05, *** indicates *p*<0.001 by one-way ANOVA with post-hoc tests. Error bars are shown as SEM (n=10 in each group). Abbreviations: AN, acinar cell necrosis; He & FN, hemorrhage and fat necrosis; Infla, inflammation; FCGBP/Fcgbp, Fc Fragment of Immunoglobulin G Binding Protein; KD, knockdown; GAPDH, glyceraldehyde-3-phosphate dehydrogenase; RNAi-AAV, adeno-associated virus packaged with small hairpin RNA; AAV, adeno-associated virus; PBS, phosphate buffer saline; ANOVA, Analysis of variance; SEM, standard error of mean.

#### Fcgbp knockdown exacerbates the severity and multi-organ injury of AP in mice

A mouse model with *Fcgbp-*knockdown (*Fcgbp-KD*) was conducted through intravenous injection of adeno-associated virus harboring a short harpin RNA targeting *Fcgbp*. Subsequently, in the colon tissue, identified as the most enriched site of Fcgbp,^24^ western-blot analysis revealed an average decrease of 41.5% in *Fcgbp* expression in *Fcgbp-KD* mice compared to the normal mice (*p*=0.002, Fig. 4B & 4C). This reduction in *Fcgbp* expression was consistently confirmed through immunohistochemistry (IHC) staining in colon tissue (Fig. 4D & 4E), lung tissue (Supplementary Fig. 6), and vessel tissue (Supplementary Fig. 7). The *Fcgbp-KD* mice, together with the mice that were injected with AAV vector or phosphate buffer saline as control, were induced to be the SAP model through intraperitoneal injection of caerulein and lipopolysaccharide. Based on pathomorphological examination, the *Fcgbp-KD* mice presented a severer morphology in pancreas tissue and lung tissue compared to the control mice (Figure 4E). Particularly, the severity of edema and destruction of alveolar structure was more pronounced in mice where *Fcgbp* was subjected to knockdown. With regards to pancreas, in line with previously established morphometry^20^ assessment for the severity of AP (Supplementary Table 14), *Fcgbp-KD* mice exhibited more severe edema (3.50 vs. 2.63, *p*=0.032), acinar cell necrosis (1.13 vs. 0.38, *p*=0.005) and inflammation (3.38 vs. 1.25, *p*<0.001) compared to control mice, suggesting the reduction of Fcgbp may cause a deleterious scenario in pancreas. In addition, the leakage of Erythrocytes into interlobular space of pancreas and pulmonary alveoli was observed in *Fcgbp-KD* mice but absent in control mice, which indicated the decrease of Fcgbp exacerbated the predisposition of vascular injury and hemorrhage (Fig. 4F & 4G).

To further explore the protective mechanism of FCGBP against SIH, we initially conducted immunofluorescence analysis on vessel tissues of mice. The results indicated that FCGBP is primarily produced by fibroblasts, as marked by α-Sma (Fig. 5A). We then performed an comprehensive examination of mRNA expression profiles of the *FCGBP*-knockdown fibroblasts compared to wild-type human vascular fibroblasts. We identified 456 genes were up-regulated and 309 genes were down-regulated in *FCGBP*-knockdown fibroblasts (Fig. 5B). Through subsequent enrichment analysis, the down-regulated gene sets were significantly enriched in terms such as “extracellular matrix”, “collagen-containing extracellular matrix”, and “external encapsulating structure” (Fig. 5C).

**Fig. 5.**
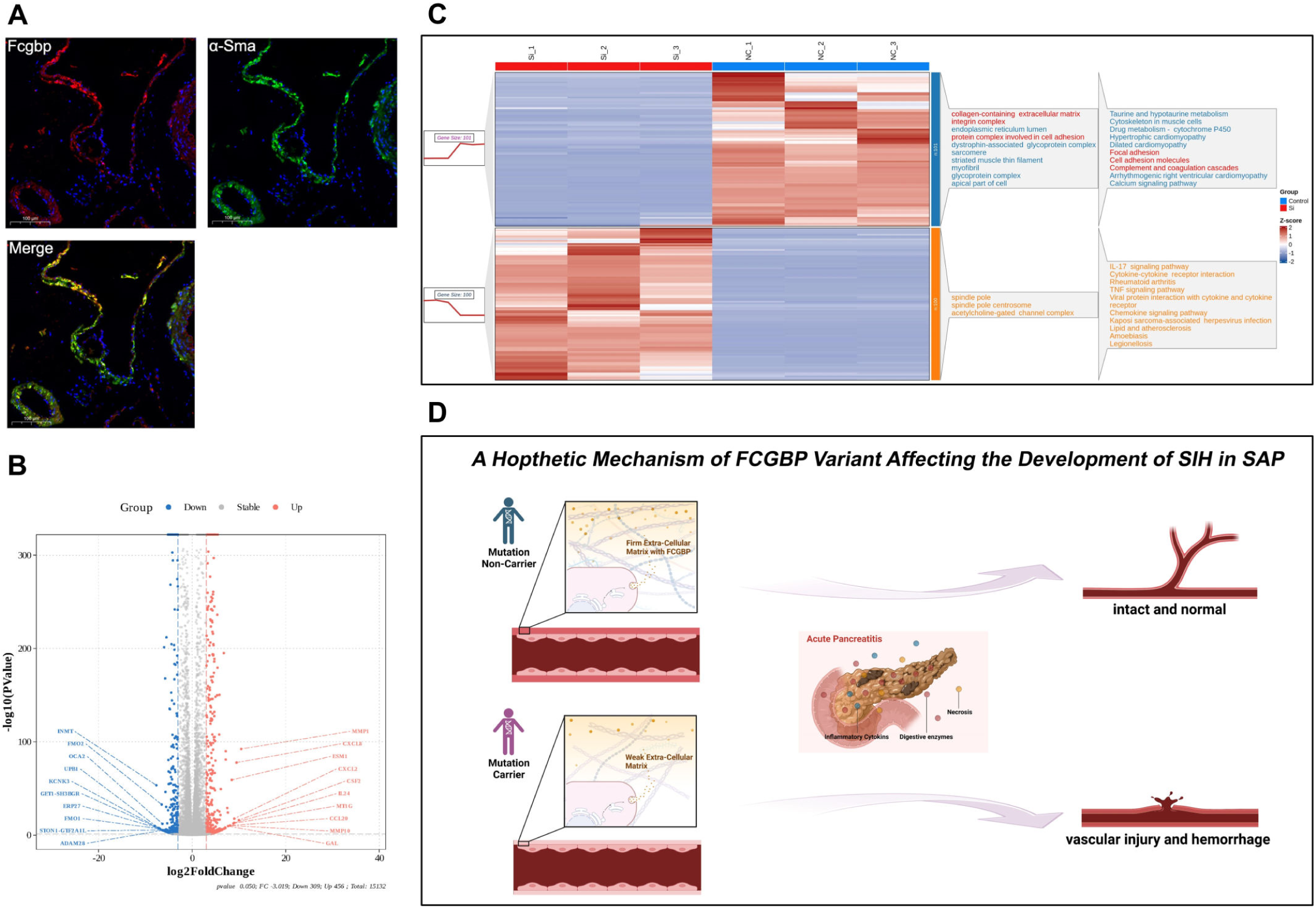
Vascular fibroblasts produce FCGBP and function as vascular wall stabilizer. (A) Representative immunofluorescence images of Fcgbp and α-Sma in vascular tissue which indicates the co-localization of Fcgbp and α-Sma (B) Volcano plot of DEGs of *FCGBP*-knockdown fibroblasts. (**C**) The top 100 up-regulated and top 101 down-regulated DEGs of FCGBP knockdown fibroblasts were presented in the heatmap plot, with enrichment items of corresponding gene sets were presented by side. (**D**) A hypothetic mechanism of *FCGBP* variants (i.e., rs1326680184) affecting the predisposition of SIH in SAP: rs1326680184 carrier suffered a weak extra cellular matrix compared with non-carrier, resulting in a higher risk of vascular injury and hemorrhage under the impact of severe acute pancreatitis. Abbreviations: FCGBP/Fcgbp, Fc Fragment of Immunoglobulin G Binding Protein; DEGs, Differentially Expressed Genes; SIH, spontaneous intraabdominal hemorrhage; SAP, severe acute pancreatitis.

#### Association between FCGBP variant of rs1326680184 and clinical characteristics

Out of 600 included participants, a total of 327 AP patients were eligible for further assessment of the association between *FCGBP* variant (e.g., rs1326680184) and clinical characteristics, based on accessibility of complete genotyping, clinical outcomes and laboratory measures. Of these included AP patients, 144 (44.0%) were rs1326680184 carriers while 183 (56.0%) were non-rs1326680184 carriers, who showed no difference on age, gender and BMI. However, the rs1326680184 carriers showed prolonged thrombin time than non-carriers (mean of 19.24s vs. 16.97s; *p*=0.010), which indicated an abnormal coagulation function affected by rs1326680184 variant occurred in *FCGBP*. In addition, the rs1326680184 carriers suffered a higher rate of mortality (11.8% vs. 4.4%; *p*=0.021), incidence of acute kidney injury (46.5% vs. 33.5%; *p*=0.023), infected pancreatic necrosis (56.2% vs. 26.9%; *p*<0.001), and critical form of AP (43.1% vs. 20.2%; *p*<0.001) (Fig. 6A; Supplementary Table 15). Those findings support the aforementioned observation that *FCGBP* variant may alter the predisposition of hemorrhage and thereby led to a poorer clinical outcomes of AP patients. Moreover, the rs1326680184 carriers tended to have a lower count of red blood cell (mean of 3.22 ×10^9^/L vs. 3.57 ×10^9^/L; *p*<0.001), hemoglobin concentration (mean of 95.36 g/L vs. 107.30 g/L; *p*<0.001) and hematocrit (mean of 29.0% vs. 30.0%; *p*=0.001) (Supplementary Table 16), providing additional evidence that phenotypes related to SIH were likely to be poorly manifested in the AP patients of presence than absence of *FCGBP* variants.

**Fig. 6.**
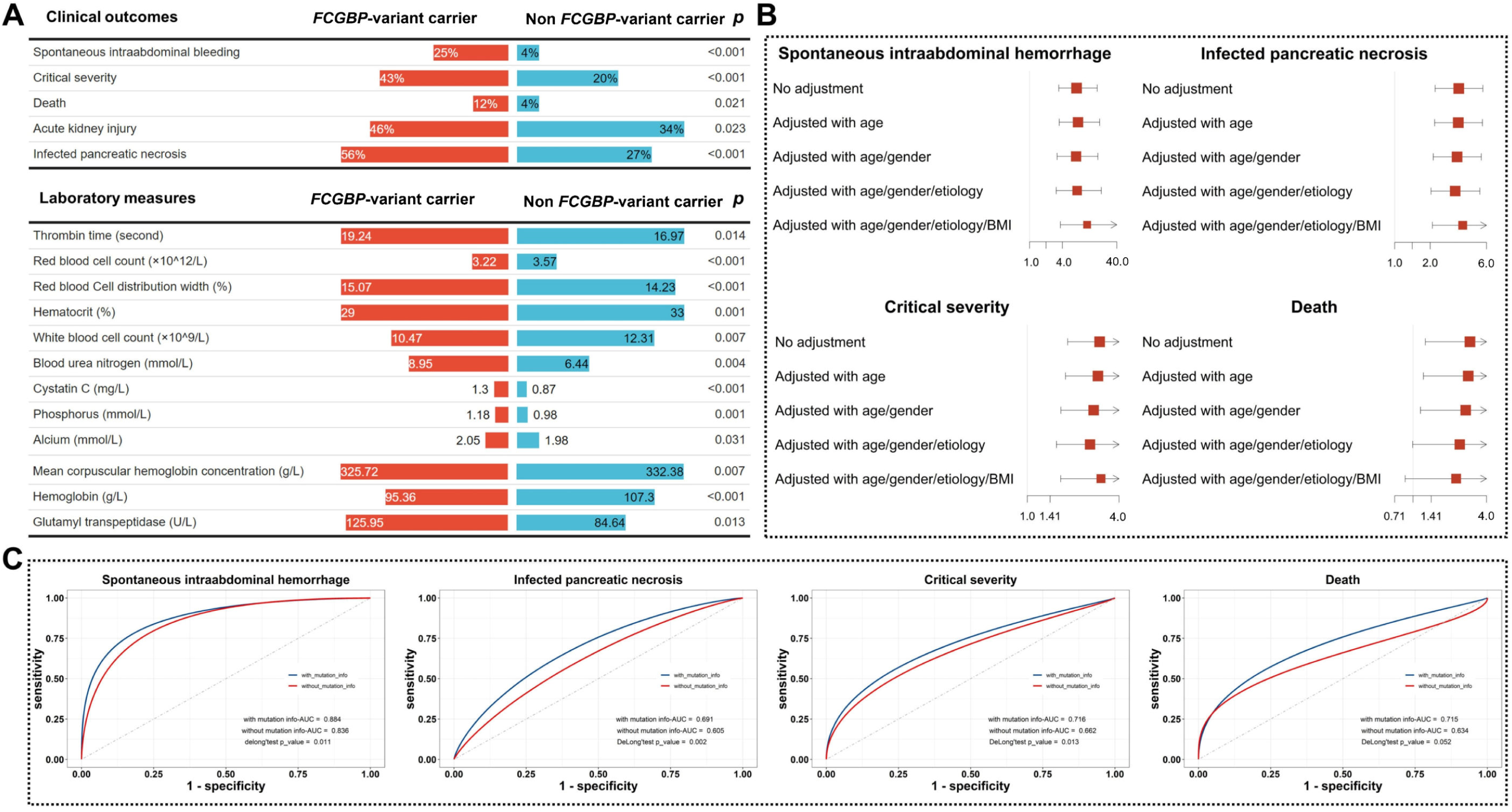
Associations between *FCGBP* variant (i.e., rs1326680184) and clinical characteristics. (**A**) a comparison of clinical characteristics at admission between rs1326680184 variant carriers and non-variant carriers. (**B**) forest plots of rs1326680184 variant with the risk of SIH, critical severity of SAP, infected pancreatic necrosis, and death. The odds ratios (ORs) were shown by solid square and horizontal lines representing the 95% confidence intervals, the adjustments were performed with age, gender, BMI, and etiology of AP. (**C**) The receiver operating characteristic curve (ROC) and diagnostic ability of demographic and rs1326680184 variant in relation to multiple clinical characteristics. The statistical comparison was conducted using DeLong’s test, which the *p*<0.05 was considered as statistical significance. Abbreviations: MAF, minor allele frequency; RNAi-AAV, adeno-associated virus packaged with small hairpin RNA; LPS, lipopolysaccharides; PBS, phosphate buffer saline; SIH, spontaneous intraabdominal hemorrhage; AP, acute pancreatitis patients; SAP, severe acute pancreatitis; ORs, odds ratios; BMI, body mass index; ROC, receiver operating characteristic curve.

To evaluate the clinically application of *FCGBP* variant rs1326680184, a standard logistic regression was performed in comparison of rs1326680184 carriers to non-carriers. The results revealed the rs1326680184 carriers with an higher risk of SIH development (OR 11.35; 95%CI: 3.66–45.81; *p*__adjust_<0.001), severity of AP (OR 3.04; 95%CI: 1.65–5.71; *p*__adjust_<0.001), and infected pancreatic necrosis (OR 3.77; 95%CI: 2.09–6.93; *p*__adjust_<0.001), respectively (Fig. 6B). Those findings were validated by using multiple models adjusted for different demographic factors. When compared to the model based on conventional demographic factors (i.e., age, gender, etiology of AP), the additional inclusion of rs1326680184 presented higher performance (i.e., area under receiver operating curve: AUROC) for diagnosing clinical outcomes respectively; SIH (0.84 vs. 0.88, *P*_-DeLong_=0.011), IPN (0.61 vs. 0.69, *P*_-DeLong_=0.002), severity of AP (0.66 vs. 0.7, *P*_-DeLong_=0.013), death (0.63 vs. 0.72, *P*_-DeLong_=0.052). These results suggest *FCGBP* variants could serve as a competent risk indicator, enhancing the diagnosis of complication and prognosis of AP with stable diagnostic ability (Fig. 6C).

## Discussion

The current study, by leveraging the largest-scale WES data of AP patients, uncovered novel insights into genetic architecture, biological mechanisms and clinical translations of a life-threatening complication (i.e., SIH), which leads to provide potential implications for critical care of AP.

By employing single-variant and gene-based analyses, this study identified the genetic variant located in *FCGBP* are enriched in patients with SAP and SIH. According to *in-silico* prediction models, the identified variant was predicted with deleterious effect and indicated an alteration of FCGBP protein levels, which was found to be associated with higher predisposition of SIH, and further related to deleterious laboratory measures and poorer prognosis in the context of AP. Those results were validated through *in-vivo* animal model, which is, the *Fcgbp* knockdown mice showed an increased severity of AP and higher predisposition of SIH, with a lower expression of *Fcgbp* in multiple organs.

Despite these findings, the underlying functions and regulatory mechanisms of *FCGBP* have not been studied in depth. In 1997, N. Harada *et al.* demonstrated that human FCGBP, which is widely expressed in systemic mucosa, exhibits mucin-like structure thereby is likely to enhance antigen trapping,^24^ which implies a pivotal role in immune and inflammatory process. Nevertheless, there is a lack of research providing robust evidence on how FCGBP functions in the human health, both in immune system and non-immune systems. While the majority of published studies were neoplastic diseases^25,26,27,28^ upon FCGBP in relation to tumor screening and prognosis, other studies has indicated that FCGBP is also involved in vascular disorders as i) Gäbel *et al.* demonstrated that alteration of genes in relation to angiogenesis including *FCGBP* were positively associated with progression and rupture of abdominal aortic aneurysms;^29^ and ii) Zhang *et al*. demonstrated *FCGBP* variant was also remarkably involved into the development and prognosis of brain arteriovenous malformation.^30^ For the first time, by using WES based on both variants and gene-set analyses, we detected and strengthened *FCGBP*-relevant variant accompanied with development of SIH in SAP patients. Specifically, we identified a frameshift deletion in *FCGBP* that may lead to an alteration of protein expression. Based on this finding, we proposed a hypothesis that the *FCGBP* variant (i.e., rs1326680184) carrier experience an alteration of FCGBP expression, increasing their susceptibility and predisposition of vascular injury and hemorrhage, which might be induced by a serious scenario (e.g., SAP; Fig. 5D). To verify this hypothesis, we used vascular tissue to identify the key cell type as the primary producer of FCGBP based on immunofluorescence analysis. We observed Fcgbp co-localized with α-Sma in the vascular tissue, suggesting vascular fibroblasts as the main producer of Fcgbp. Therefore, we cultured human vascular fibroblasts with knocking down FCGBP, followed by mRNA expression profiling. We found differentially regulated genes were enriched in the function for vascular stability. Such findings support our hypothesis that down-regulated *FCGBP* expression could destabilize the vascular wall, and lead to predisposition of vascular injury in the context of SAP. However, due to the concealed protein structure of FCGBP, the exploration for details of underlying mechanistic pathways of FCGBP on SIH remains constrained, further substantial studies with in-depth molecular experiments are warrant.

To further explore the translational significance of the above findings, we developed a clinical classification model that demonstrated improved diagnostic capacity with the inclusion of information of *FCGBP* variant, not only for SIH but also for IPN, severity and mortality of AP. The FCGBP alteration could therefore, be considered as an indicator or therapeutic target for SAP and extensive critical diseases, which could enhance the precision of prognostic assessments, helping to stratify patients for specific treatments and clinical care.

To our knowledge, this is the first attempt to discover novel genetic associations by WES in a population of patients with AP. Here we identified strong associations of exonic variant located in FCGBP with SIH, which interacted with other complications and further related to the severity and mortality of AP. Those findings might direct the development of highly needed therapies to prevent life-threatening complications in the early phase of acute pancreatitis, thereby facilitating a more effective strategy for AP prognosis and critical care.

Here are some limitations that require acknowledgement in our study; first, although our cohort consists of a rare population of severely AP patients with SIH, the sample size of our primary discovery was relatively small in terms of genetic research. By applying the multi-stage analyses and experimental verification, we were able to limit our sample size and still be able to detect true differences between the phenotypes; second, while this study represented large-scale AP patients transferred from multi centers across China, our analyses were still limited by lack of external validation. The correlation between genetic alterations and clinicopathological characteristics, should still be fully validated in an independent population; third, because of the retrospective study design, residual bias could not be fully eliminated. Nevertheless, through stringent clinical assessment by well-trained doctors and precise laboratory measures, the results affected by residual bias would be minimized. The retrospective nature of such study also resulted in that some of the sample quality failed to fully meet the criteria of sequencing, that is, the bias of WES quality could not be completely ruled out. In addition, due to the technical and annotated limit of WES, the structural variants (e.g., copy number variants) were difficult to accurately capture; fourth, the current study was conducted in a predominantly Chinese population, therefore, findings should be implemented with caution when generalized to other ethnicities; fifth, despite we carefully matched our population, it is possible that other unmeasured factors were responsible for the observed difference in allele frequency and alter our results; sixth, even though the *Fcgbp-KD* mouse model provided sufficient *in-vivo* verification for populational observation of *FCGBP* variant, the use of *Fcgbp* knockout (*Fcgbp^-/-^*) mouse model might augment the experimental evidence. However, the necessity of using *Fcgbp* knockout (*Fcgbp^-/-^*) mouse model still remained dispute with consideration of the against on the uncertainty of function of *FCGBP* variant and the mitigation of suffering for experimental animals; seventh, due to the unknown *FCGBP* gene structure and its protein structure, the demonstration in terms of the formality caused by *FCGBP* variant is impeded, thereby future structural biological research is warrant. finally, while we are only beginning to dissect the genomic architecture that drives complication (i.e., SIH) in AP with a hypothesized mechanism, this study adds considerable insights and provides leads for further functional analyses or targets for therapies in the context of AP, and deserves to expand to other complications.

## Data Availability

The original data and codes used for the current study are available upon reasonable request. The raw sequencing data reported in this study have been being deposited and will be publicly available after this paper is published. All data are provided within the article.

https://ngdc.cncb.ac.cn/gsa-human/

## Ethics statements

### Patient consent for publication

All participants were informed about the purpose of study during the admission interview, with consent prior to enrolling in the study.

### Ethics approval

The study design and conduction complied with all relevant regulations regarding the human participants and was in accordance with the criteria set by the Declaration of Helsinki. The study’s protocol was approved by the Ethics Committee of Jinling Hospital, Nanjing, China (ref. No. 2021NZKY-042-01).

All animal experiment procedures followed Chinese Animal Welfare Act, the Guidance for Animal Experimentation of Southeast University. Efforts were made to minimize the number of animals used and reduce their suffering (ref. No. 2022DZGKJDWLS-0059).

## Declaration of interests

The authors declare no competing interests.

## Author contributions

Conceived and designed the study: Q.Y.T, E.Y.Y; supervision: E.Y.Y, W.Q.L; conducted data analyses and interpretation and drafted the manuscript: Q.Y.T; data curation: Q.Y.T, Y.P.H, Q.Y; conducted experiments: Y.P.H, Q.Y.T; critical revision of the manuscript: E.Y.Y, W.Q.L, L.K, Y.X.L, J.Z, J.Z.Z, J.Y, H.B.H., G.L, B.Q.L, All authors read and approved the final manuscript.

## Acknowledgments

This research was funded by the National Natural Science Foundation of China (8207035023, 82204033), Natural Science Foundation of Jiangsu Province (BK20220826). Dr. E.YW.Yu is recipient of the Zhishan Young Scholar Award at the Southeast University (2242023R40031). The authors acknowledge that certain figures were created, adapted and exported from BioRender.com (2023). Retrieved from https://app.biorender.com/biorender-templates.

## Graphical Abstract

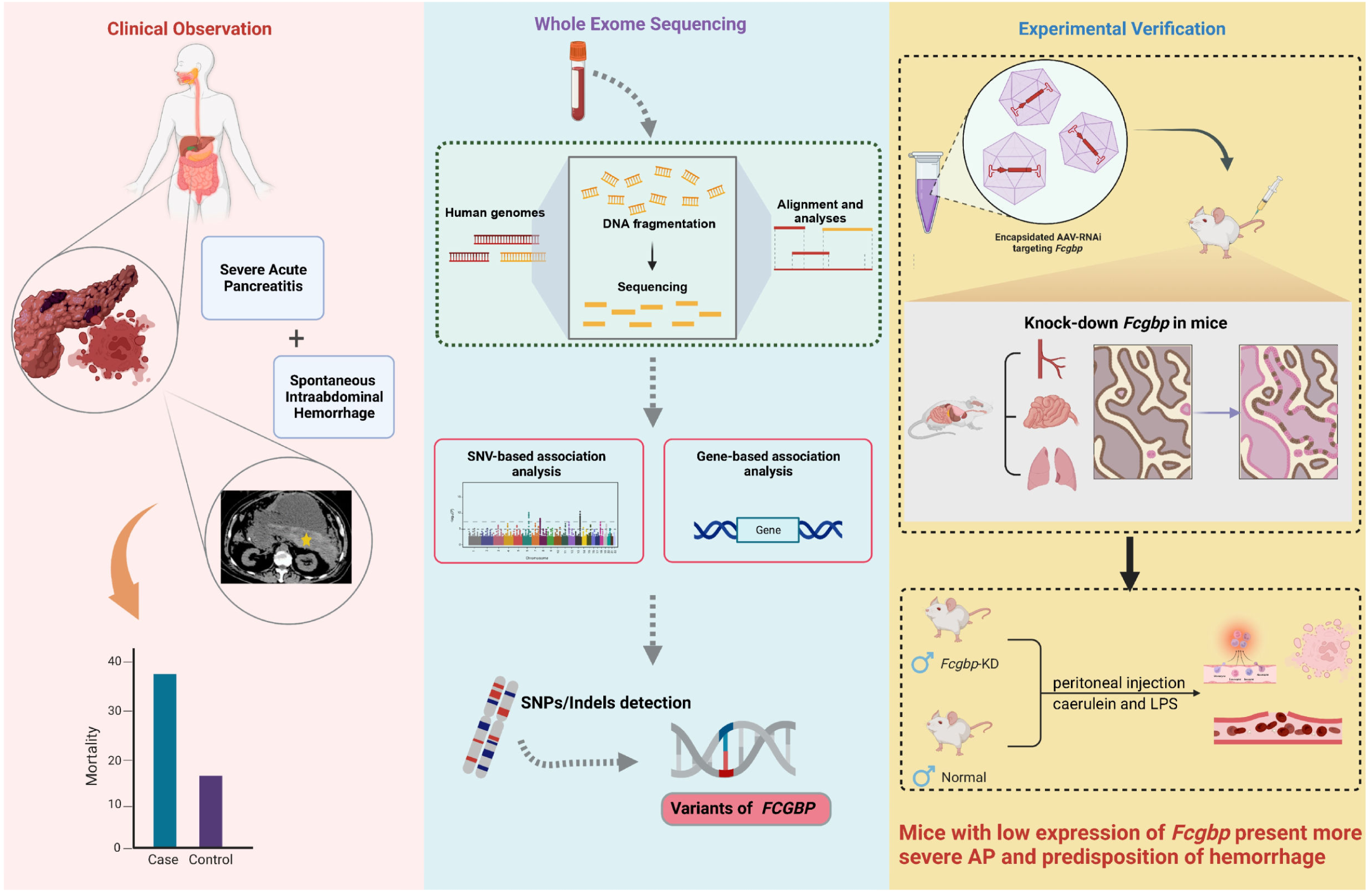

## Notes

### Competing Interest Statement

The authors have declared no competing interest.

### Author Declarations

Ethics Committee of Jinling Hospital, Nanjing, China gave ethical approval for this work.(No.2021NZKY-042-01,No.2022DZGKJDWLS-0059)

