## Supplemental methods, figures and tables for "Whole Exome Sequencing Reveals *FCGBP* Variant Associated with Spontaneous Intraabdominal Hemorrhage in Severe Acute Pancreatitis"

This appendix formed part of the original submission.

^*^Wei-Qin Li, M.D.

Mailing address: No. 305 Zhongshan East Road, Nanjing 210002, China.

&

^**^Evan Yi-Wen Yu, Ph.D.

Phone number: +31 06 13155188

Mailing address: 87 Ding Jiaqiao Rd, Nanjing 210009, China

ORCID: <https://orcid.org/0000-0001-7825-5087>

**CONTENT OF SUPPLEMENTARY MATERIALS**

**Supplementary Methods**

**Supplementary Table 1** Designation of Fcgbp-targeted small hairpin RNA

**Supplementary Table 2** Quality control measures and summary sequence statistics of whole exome sequencing

**Supplementary Table 3** Genomic coordinates of candidate variants and allele frequencies retrieved from 1000 Genome Project

**Supplementary Table 4** Effects of candidate variants on protein function based on combined annotation dependent depletion (CADD) score

**Supplementary Table 5** Effects of candidate variants on protein function based on Polymorphism Phenotyping (PolyPhen)

**Supplementary Table 6** Effects of candidate variants on protein function based on Sorting Intolerant from Tolerant algorithm (SIFT)

**Supplementary Table 7** Associations between identified candidate variants and clinical phenotypes based on ClinVar database

**Supplementary Table 8** Prior evidence of identified candidate variants based on published GWASs

**Supplementary Table 9** Epigenomic information for candidate variants based on Roadmap database

**Supplementary Table 10** Indicated mechanistic pathway for identified candidate variants

**Supplementary Table 11** Estimated probability score for conserved region of the candidate variants

**Supplementary Table 12** CpG annotation of candidate variants

**Supplementary Table 13** The genes identified to be associated with SIH based on SKAT analysis

**Supplementary Table 14** Morphometry quantification of the severity of acute pancreatitis

**Supplementary Table 15** Comparison of distributions for clinical outcomes between rs1326680184 carriers and non rs1326680184 carriers

**Supplementary Table 16** Comparison of distributions for laboratory measures between rs1326680184 carriers and non rs1326680184 carriers

**Supplementary Fig. 1** Pathology examination of lung and pancreas tissue of mice

**Supplementary Fig. 2** Assays of blood MCP1, TNFα and IL-6 from mice induced as AP and normal healthy mice

**Supplementary Fig. 3** Quantile-quantile plot for the GWAS analysis of WES and SIH

**Supplementary Fig. 4** Manhattan plot of genome-wide association analysis for exonic variants in all autosomal chromosomes

**Supplementary Fig. 5** Assay of serum FCGBP in patients with or without SIH

**Supplementary Fig. 6** Immunohistochemical analysis of *Fcgbp* expression in lung tissue in mice

**Supplementary Fig. 7** Immunohistochemical analysis of *Fcgbp* expression in vessel tissue in mice

**Supplementary methods**

***Quality control of DNA sample in whole exome sequencing***

The genomic DNA extracted from peripheral blood was undertaken quality controls by: i) analysis of the degree of degradation and pollution of RNA with agarose gel electrophoresis; ii) test of the DNA purity with Nanodrop (Thermo Fisher Scientific, Waltham, MA), which is a spectrophotometer that enables highly accurate analyses; iii) meticulous quantification of DNA concentration with Qubit 3.0 (Thermo Fisher Scientific, Waltham, MA).

***Sequencing data processing including variants calling and quality control***

Sample reads mapping, variant calling, annotation, and quality control were performed using a pre-planned protocol, which were conducted in all participants irrespective of the outcomes (i.e., cases or controls). Quality control for raw FASTQ data was performed using software FastQc (version 0.11.9) to summarize statistics such as sequence count, sequence quality, GC content, adapter content, base N content, sequence length distribution, sequence duplication levels, and overrepresented sequence. MultiQC (version 1.14) was used to aggregate per-sample results into a single report. The sequence matched to the sequencing adapters and low-quality ends of reads (the Phred-scale quality score of base<20) were removed with Trim-galore (version 0.6.7). The trimmed sequences were then interatly aligned to the human genome reference version hg38 (http://genome.ucsc.edu/) using Burrows-Wheeler Aligner (BWA) software (version 0.7.8-r455) with Maximum Entropy Method (BWA-MEM) algorithm; the output SAM files were transformed to binary format and indexed with SAMtools (version 1.9). Base score recalibration, local realignment for indel (insertion or deletion) detection, and duplication mark were performed with Genome Analysis Tool Kit (GATK, version 4.2.2.0, Broad Institute). The joint variant calling of germline variants was performed following GATK workflow for case group and control group, respectively, in which per-sample intermediate data were gathered and then produced a set of joint-called variants to enhance the sensitive detection of variants.

To validate the accuracy of variant calling obtained above, we additionally used a deep learning-based variant calling method, i.e., DeepVariant (version 1.3.0), to accomplish an independent procedure of variant calling, in which the two workflows (i.e., GATK and DeepVariant) performed largely equal on mapping the exome-wide variants.

The variants, with either missing rate >0.50, minimal frequency of alternative alleles in case group <3, Phred-scaled quality value <30, or disqualified variant quality score recalibration (VQSR) (i.e., a new variant quality score named VQSLOD provides a continuous estimate of the probability that each variant is true and a threshold of 99.00 was applied), were removed using VCFtools (version 0.1.16).

***Functional annotation***

An in-silico approach through SNPnexus (<https://www.snp-nexus.org/v4/>), ANNOVAR (http://www.openbioinformatics.org/annovar), and RegulomeDB (https://beta.regulomedb.org/regulome-search/) was used to annotate the exonic variants. The allele frequencies of a given variant in large populations (particularly East Asians) were aligned with the Exome Aggregation Consortium database and 1000 Genomes Project database. Variants were defined as “uncommon” if they were present in <1% of population in the reference databases. However, it is important to note that some variants, such as rs1326680184, did not have available allele frequency information in the genome reference database and were consequently not filtered out from further analysis. Genetic variants predicted to be deleterious were identified if they disrupt the protein-coding sequence (refer to nonsynonymous, stop-gained, start-loss, frameshift, or canonical splicing-site alteration). In addition, to examine predicted functional impact, particularly for the frameshit mutations that could not be annotated by neither Polymorphism Phenotyping (PolyPhen) nor Sorting Intolerant from Tolerant algorithm (SIFT) designed for single-nucleotide polymorphisms (SNPs), a combined annotation dependent depletion (CADD) method was annotated to the variants (Phred scores >20 predicted as deleterious, <https://cadd.gs.washington.edu/score>).

***ELISA***

Concentrations of human serum FCGBP were detected with a Human FCGBP ELISA Kit (RK01369, ABclonal, China) following the manufacturer’s instructions, and compared between FCGBP variant carriers and non-carriers by Student’s t test with Welch’s correction. Mouse serum levels of MCP1, IL-6 and TNFα were measured using MCP1, IL-6 and TNFα ELISA kits (RK00381, RK00008, RK00027, ABclonal, China), which the differences between AP mice and control mice were compared using Man-Whitney U test.

***Western-blot***

Briefly, tissues from mice were lysed in RIPA lysis buffer, and total proteins were extracted. Concentrations of proteins were measured using the BCA protein assay kit (Pierce, Thermo Fisher Scientific) following the commercial instructions. Equivalent protein (approximately 30 μg/sample) was separated by SDS-PAGE gel and then transferred to PVDF membranes. Then the membranes were blocked for 1 hour in 5% nonfat milk. Afterwards, membranes were incubated with primary antibodies against GAPDH (1:1,000 dilution; Sigma-Aldrich), Fcgbp (1:1,000 dilution; Bioss) overnight at 4°C. Finally, HRP-conjugated secondary antibodies were applied for 1 hour. The proteins were detected and analyzed by an ECL Plus chemiluminescence imaging system (Tanon). The quantified results of Western-blot were compared between Fcgbp-KD mice and control mice using Man-Whitney U test.

**Supplementary Table 1(a) The designated AAV-RNAi targeting *Fcgbp***

| **Target sequence** | **GC%** |
| --- | --- |
| TCGTGATTGAAACCGACTTTG | 0.4211 |

**Supplementary Table 1(b) The synthesized oligo information**

|  | **5’ sequence** | **Stem sequence** | **Loop** | **Stem sequence** | **3’ sequence** |
| --- | --- | --- | --- | --- | --- |
| Fcgbp-RNAi (115174-1)-a | ACCGG | TCGTGATTGAAACCGACTTTG | TTCAAGAGA | CAAAGTCGGTTTCAATCACGA | TTTTT |
| Fcgbp-RNAi (115174-1)-b | TCTAAAAAA | TCGTGATTGAAACCGACTTTG | TCTCTTGAA | CAAAGTCGGTTTCAATCACGA | C |

For *Fcgbp*-target AAV-RNAi packaging, the designed shRNA primers were inserted into the vector. The titer of AAV were 3.36×10^13^ viral genome copies per ml, and a total of 5.00×10^11^ viral genome copy was infused through intravenous injection per mouse. The atlas of vector is accessible at <http://www.genechem.com.cn/service/index.php?ac=gene&at=vector_search&keyword>

Abbreviations: AAV, adeno-associated virus; AAV-RNAi, adeno-associated virus harboring short interfering RNA.

**Supplementary Table 2 Quality control measures and summary sequence statistics of the whole exome sequencing.**

| **ID** | **I7 index** | **I5 index** | **Reads** | **Length** | **amount of data (G)** | **%Q20** | **%Q30** | **%GC** |
| --- | --- | --- | --- | --- | --- | --- | --- | --- |
| 1 | CTGAAGCT | AGGCGAAG | 40717395.00 | 150 | 12.22 | 98.05 | 94.68 | 51.33 |
| 2 | CGGCTATG | TATAGCCT | 38378429.00 | 150 | 11.51 | 98.08 | 94.78 | 51.02 |
| 3 | CGGCTATG | GGCTCTGA | 35575748.00 | 150 | 10.67 | 98.08 | 94.78 | 51.05 |
| 4 | CGGCTATG | AGGCGAAG | 45746403.00 | 150 | 13.72 | 98.07 | 94.77 | 51.28 |
| 5 | CTGAAGCT | TAATCTTA | 50557030.00 | 150 | 15.17 | 98.09 | 94.81 | 51.79 |
| 6 | TCCGCGAA | GTACTGAC | 51582329.00 | 150 | 15.47 | 98.06 | 94.75 | 51.05 |
| 7 | CTGAAGCT | CAGGACGT | 41058312.00 | 150 | 12.32 | 98.02 | 94.63 | 51.36 |
| 8 | CTGAAGCT | GTACTGAC | 37961418.00 | 150 | 11.39 | 98.0 | 94.59 | 51.2 |
| 9 | TAATGCGC | TATAGCCT | 36986728.00 | 150 | 11.1 | 98.03 | 94.65 | 51.03 |
| 10 | TAATGCGC | ATAGAGGC | 38651255.00 | 150 | 11.6 | 98.02 | 94.59 | 50.7 |
| 11 | TAATGCGC | CCTATCCT | 39552701.00 | 150 | 11.87 | 98.0 | 94.55 | 50.84 |
| 12 | TAATGCGC | GGCTCTGA | 37762436.00 | 150 | 11.33 | 98.03 | 94.62 | 50.76 |
| 13 | TAATGCGC | TAATCTTA | 40080249.00 | 150 | 12.02 | 98.11 | 94.81 | 51.04 |
| 14 | TAATGCGC | CAGGACGT | 42812851.00 | 150 | 12.84 | 98.0 | 94.59 | 51.45 |
| 15 | TAATGCGC | AGGCGAAG | 38041403.00 | 150 | 11.41 | 98.04 | 94.62 | 50.64 |
| 16 | TAATGCGC | GTACTGAC | 37645964.00 | 150 | 11.29 | 97.21 | 92.53 | 50.88 |
| 17 | CGGCTATG | ATAGAGGC | 36850441.00 | 150 | 11.06 | 97.16 | 92.49 | 50.58 |
| 18 | CGGCTATG | CCTATCCT | 39242995.00 | 150 | 11.77 | 97.28 | 92.73 | 50.66 |
| 19 | CGGCTATG | TAATCTTA | 44505581.00 | 150 | 13.35 | 97.29 | 92.75 | 50.33 |
| 20 | CGGCTATG | CAGGACGT | 44405141.00 | 150 | 13.32 | 97.14 | 92.44 | 50.34 |
| 21 | CGGCTATG | GTACTGAC | 40991736.00 | 150 | 12.3 | 97.14 | 92.44 | 50.34 |
| 22 | TCCGCGAA | ATAGAGGC | 46366009.00 | 150 | 13.91 | 97.05 | 92.27 | 49.96 |
| 23 | TCCGCGAA | CCTATCCT | 45472952.00 | 150 | 13.64 | 97.06 | 92.28 | 50.06 |
| 24 | TCCGCGAA | CAGGACGT | 50051875.00 | 150 | 15.02 | 97.06 | 92.31 | 50.06 |
| 25 | TCCGCGAA | GGCTCTGA | 40445348.00 | 150 | 12.13 | 97.03 | 92.22 | 50.2 |
| 26 | TCCGCGAA | AGGCGAAG | 43527498.00 | 150 | 13.06 | 97.13 | 92.41 | 49.78 |
| 27 | TCCGCGAA | TAATCTTA | 44622233.00 | 150 | 13.39 | 97.15 | 92.47 | 49.89 |
| 28 | TCCGCGAA | TATAGCCT | 39120397.00 | 150 | 11.74 | 97.0 | 92.2 | 50.47 |
| 29 | TCTCGCGC | ATAGAGGC | 43958670.00 | 150 | 13.19 | 97.01 | 92.14 | 48.61 |
| 30 | TCTCGCGC | CCTATCCT | 46755872.00 | 150 | 14.03 | 97.03 | 92.19 | 49.01 |
| 31 | TCTCGCGC | TATAGCCT | 38179657.00 | 150 | 11.45 | 96.98 | 92.14 | 48.94 |
| 32 | TCTCGCGC | GGCTCTGA | 42321689.00 | 150 | 12.7 | 97.01 | 92.15 | 48.58 |
| 33 | TCTCGCGC | GTACTGAC | 40206625.00 | 150 | 12.06 | 97.12 | 92.38 | 48.52 |
| 34 | TCTCGCGC | AGGCGAAG | 35660704.00 | 150 | 10.7 | 97.14 | 92.41 | 48.52 |
| 35 | TCTCGCGC | TAATCTTA | 37001564.00 | 150 | 11.1 | 97.13 | 92.41 | 48.73 |
| 36 | TCTCGCGC | CAGGACGT | 38836488.00 | 150 | 11.65 | 97.01 | 92.15 | 48.73 |
| 37 | AGCGATAG | TATAGCCT | 41457473.00 | 150 | 12.44 | 97.14 | 92.53 | 50.91 |
| 38 | AGCGATAG | ATAGAGGC | 38692714.00 | 150 | 11.61 | 97.18 | 92.56 | 50.83 |
| 39 | AGCGATAG | GGCTCTGA | 37642683.00 | 150 | 11.29 | 97.15 | 92.5 | 50.77 |
| 40 | AGCGATAG | CCTATCCT | 38679561.00 | 150 | 11.6 | 97.2 | 92.63 | 50.73 |
| 41 | AGCGATAG | AGGCGAAG | 41239636.00 | 150 | 12.37 | 97.28 | 92.78 | 50.79 |
| 42 | AGCGATAG | TAATCTTA | 44083619.00 | 150 | 13.23 | 97.22 | 92.72 | 51.12 |

Sequencing data were produced on Illumina NovaSeq 6000 System, with a paired-end strategy (2×150 base-pair) and transformed to FASTQ format. Q20 and Q30 refer to quality scores that measure the probability that a base is called incorrectly. A quality score of 20 (Q20) represents an error rate of 1 in 100 (meaning every 100 bp sequencing read may contain an error), with a corresponding call accuracy of 99%. A quality score of 30 (Q30) represents an error rate of 1 in 1000 (meaning every 1000 bp sequencing read may contain an error). The percentage of GC represents Count (G + C)/Count (A + T + G + C) * 100%.

Please see **Supplementary Table 3-12** in the additional Excel file.

**Supplementary Table 13 The genes identified to be associated with SIH based on SKAT analysis.**

| **Gene** | ***p-*value** |
| --- | --- |
| *FCGBP* | 1.31×10^-7^ |
| *PABPC3* | 3.92×10^-9^ |
| *MYCN* | 8.95×10^-6^ |
| *PLIN4* | 9.00×10^-6^ |
| *MAP1A* | 0.00015 |
| *RAD51B* | 0.000424 |
| *CREB3L1* | 0.000659 |
| *MYH14* | 0.000869 |
| *MUC20* | 0.001164 |
| *ZNF626* | 0.00159 |
| *EBF4* | 0.00258 |
| *KCNJ18* | 0.003938 |
| *MST1L* | 0.004935 |
| *PEX5* | 0.005769 |
| *FAM160A2* | 0.006143 |
| *MUC3A* | 0.008796 |
| *PNKD* | 0.009227 |

SKAT analysis uses a kernel function to compute a similarity measure between each pair of genetic variants based on their allele frequencies and genetic distances. This kernel matrix is then used to calculate a test statistic that determines the degree of association between the genetic variants and the phenotype. Only the genes with *p*<0.01 were reported.

Abbreviation: SKAT: Sequence Kernel Association Test.

**Supplementary Table 14 Morphometry quantification of the severity of acute pancreatitis.^1^**

| **Edema** |  |
| --- | --- |
| 0 | Absence |
| 0.5 | Focal expansion of interlobar septa |
| 1 | Diffuse expansion of interlobar septa |
| 1.5 | Same as 1+focal expansion of interlobular septa |
| 2 | Same as 1+diffuse expansion of interlobular septa |
| 2.5 | Same as 2+focal expansion of interacinar septa |
| 3 | Same as 2+diffuse expansion of interacinar septa |
| 3.5 | Same as 3+focal expansion of intercellular spaces |
| 4 | Same as 3+diffuse expansion of intercellular spaces |
| **Acinar necrosis** |  |
| 0 | Absence |
| 0.5 | Focal occurrence of 1-4 necrotic cells/HPF |
| 1 | Diffuse occurrence of 1-4 necrotic cells/HPF |
| 1.5 | Same as 1+focal occurrence of 5-10 necrotic cells/HPF |
| 2 | Diffuse occurrence of 5-10 necrotic cells/HPF |
| 2.5 | Same as 2+focal occurrence of 11-16 necrotic cells/HPF |
| 3 | Diffuse occurrence of 11-16 necrotic cells/HPF (foci of confluent necrosis) |
| 3.5 | Same as 3+focal occurrence of >16 necrotic cells/HPF |
| 4 | >16 necrotic cells/HPF (extensive confluent necrosis) |
| **Hemorrhage and fat necrosis** |  |
| 0 | Absence |
| 0.5 | 1 focus |
| 1 | 2 foci |
| 1.5 | 3 foci |
| 2 | 4 foci |
| 2.5 | 5 foci |
| 3 | 6 foci |
| 3.5 | 7 foci |
| 4 | ≥8 foci |
| **Inflammation and perivascular infiltration** |  |
| 0 | 0-1 intralobular or perivascular leukocytes/HPF |
| 0.5 | 2-5 intralobular or perivascular leukocytes/HPF |
| 1 | 6-10 intralobular or perivascular leukocytes/HPF |
| 1.5 | 11-15 intralobular or perivascular leukocytes/HPF |
| 2 | 16-20 intralobular or perivascular leukocytes/HPF |
| 2.5 | 21-25 intralobular or perivascular leukocytes/HPF |
| 3 | 26-30 intralobular or perivascular leukocytes/HPF |
| 3.5 | >30 leukocytes/HPF or focal micro abscess |
| 4 | >35 leukocytes/HPF or confluent micro abscess |

The morphological examination, e.g., edema area and grade, acinar cell necrosis, inflammatory reaction, adipose necrosis, and hemorrhage, was assessed to determine the severity of AP for each group of mice (n=10 for each group) by two independent and blinded investigators, using a previously reported morphometry guideline.^1^

Abbreviation: HPF, high power field; AP, acute pancreatitis.

**Supplementary Table 15 Comparison of distributions for clinical outcomes between *FCGBP* variants (e.g., rs1326680184) carriers and non-rs1326680184 carriers.**

| **Characteristics** | **Overall**  **n=327 (%)** | **rs1326680184 carrier**  **n=144 (%)** | **Non-rs1326680184 carrier**  **n=183 (%)** | ***p*-value** |
| --- | --- | --- | --- | --- |
| **SIH** | 44 (13.5) | 36 (25.0) | 8 (4.4) | <0.001 |
| **Etiology** |  |  |  |  |
| Alcohol | 3 (0.9) | 2 (1.4) | 1 (0.5) | 0.133 |
| Biliary | 34 (10.4) | 21 (14.6) | 13 (7.1) |  |
| HTG | 280 (85.6) | 117 (81.2) | 163 (89.1) |  |
| others | 10 (3.1) | 4 (2.8) | 6 (3.3) |  |
| **Severity** |  |  |  |  |
| Mild | 46 (14.1) | 16 (11.1) | 30 (16.4) | <0.001 |
| Moderate | 98 (30.0) | 34 (23.6) | 64 (35.0) |  |
| Severe | 84 (25.7) | 32 (22.2) | 52 (28.4) |  |
| Critical | 99 (30.3) | 62 (43.1) | 37 (20.2) |  |
| **Death** | 25 (7.6) | 17 (11.8) | 8 (4.4) | 0.021 |
| **ARDS** | 150 (45.9) | 70 (48.6) | 80 (43.7) | 0.441 |
| **AKI** | 128 (39.3) | 67 (46.5) | 61 (33.5) | 0.023 |
| **IPN** | 130 (39.9) | 81 (56.2) | 49 (26.9) | <0.001 |
| **IAH** | 25 (7.7) | 11 (7.6) | 14 (7.7) | 0.999 |
| **Liver dysfunction** | 145 (44.5) | 65 (45.1) | 80 (44.0) | 0.919 |

*p*-values were derived from *Chi-square* test for categorical variables.

Abbreviations: SIH, Spontaneous Intrabdominal Hemorrhage; ARDS, Acute Respiratory Distress Syndrome; AKI, Acute Kidney Injury; IPN,Infected Pancreatic Necrosis; IAH, Intra-Abdominal Hypertension; HTG, Hypertriglyceridemia.

**Supplementary Table 16 Comparison of distributions for laboratory measures between *FCGBP* variants (e.g., rs1326680184) carriers and non-rs1326680184 carriers.**

| **Laboratory measures** | **Overall** n=327 | **FCGBP-mutation carrier** n=144 | **Non FCGBP-mutation** carrier n=183 | ***p*** |
| --- | --- | --- | --- | --- |
| **Prothrombin time (second)** | 13.70 (12.70, 14.90) | 13.50 (12.60, 14.67) | 13.90 (12.80, 15.20) | 0.156 |
| **Thrombin time (second)** | 15.70 (14.80, 17.75) | 15.50 (14.40, 17.40) | 16.30 (15.12, 18.62) | 0.014 |
| **Activated partial thromboplastin time (second)** | 31.00 (27.30, 35.80) | 31.00 (27.30, 35.60) | 30.85 (27.17, 37.08) | 0.053 |
| **Antithrombin Ⅲ (%)** | 65.50 (55.18, 79.55) | 64.55 (55.32, 78.12) | 66.95 (55.13, 81.45) | 0.26 |
| **INR** | 1.19 (1.11, 1.30) | 1.18 (1.10, 1.27) | 1.21 (1.11, 1.32) | 0.291 |
| **D Dimer (mg/L)** | 4.62 (2.60, 8.02) | 4.70 (2.56, 8.41) | 4.54 (2.79, 7.72) | 0.93 |
| **Neutrophil count (×10^9^/L)** | 7.89 (5.26, 10.84) | 8.32 (5.28, 12.18) | 7.57 (5.15, 9.88) | 0.703 |
| **Platelet count (×10^9^/L)** | 203.00 (135.25, 287.75) | 214.00 (146.50, 297.50) | 184.00 (124.50, 269.00) | 0.094 |
| **Platelet distribution width (%)** | 13.40 (11.25, 16.30) | 13.00 (11.05, 15.80) | 13.50 (11.40, 16.72) | 0.394 |
| **Mean platelet volume (fL)** | 11.00 (10.10, 11.80) | 11.00 (10.10, 11.70) | 11.00 (10.15, 11.90) | 0.356 |
| **Eosinophil count (×10^9^/L)** | 0.05 (0.01, 0.09) | 0.06 (0.02, 0.09) | 0.04 (0.01, 0.09) | 0.171 |
| **Platelet hematocrit** | 0.21 (0.15, 0.30) | 0.23 (0.16, 0.31) | 0.21 (0.14, 0.28) | 0.056 |
| **Mean corpuscular hemoglobin concentration (g/L)** | 328.00 (318.00, 338.00) | 331.00 (320.00, 342.00) | 325.00 (314.00, 336.00) | 0.007 |
| **Mean corpuscular hemoglobin (pg)** | 29.90 (28.70, 31.00) | 30.00 (28.90, 31.20) | 29.80 (28.60, 30.92) | 0.179 |
| **Mean corpuscular volume (fL)** | 91.30 (88.00, 93.93) | 91.10 (88.00, 93.50) | 91.60 (87.97, 94.70) | 0.257 |
| **Lymphocyte count (×10^9/^L)** | 0.98 (0.69, 1.30) | 1.07 (0.80, 1.29) | 0.87 (0.60, 1.29) | 0.083 |
| **Red blood cell count (×10^12^/L)** | 3.33 (2.73, 3.94) | 3.47 (2.82, 4.18) | 3.18 (2.58, 3.73) | <0.001 |
| **Red blood Cell distribution width (%)** | 14.25 (13.30, 15.60) | 14.00 (13.20, 14.90) | 14.60 (13.80, 16.30) | <0.001 |
| **Hemoglobin (g/L)** | 96.50 (81.00, 119.75) | 101.00 (84.00, 125.00) | 93.00 (76.00, 110.00) | <0.001 |
| **Hematocrit** | 0.30 (0.25, 0.36) | 0.32 (0.26, 0.38) | 0.29 (0.25, 0.33) | 0.001 |
| **Monocytes count (×10^9^/L)** | 0.55 (0.35, 0.77) | 0.58 (0.41, 0.77) | 0.52 (0.33, 0.76) | 0.614 |
| **White blood cell count (×10^9^/L)** | 10.46 (7.50, 14.10) | 11.29 (8.25, 14.93) | 9.08 (6.86, 12.07) | 0.007 |
| **Fibrinogen (g/L)** | 4.98 (3.84, 7.00) | 4.97 (4.00, 7.00) | 5.01 (3.58, 6.97) | 0.521 |
| **Fibrinogen degradation products (μg/L)** | 16.60 (10.70, 30.10) | 17.20 (10.80, 29.90) | 15.90 (10.70, 30.10) | 0.91 |
| **Procalcitonin (μg/L)** | 0.63 (0.22, 2.45) | 0.57 (0.22, 1.97) | 0.83 (0.22, 3.25) | 0.282 |
| **Interleukin 6 (ng/L)** | 87.40 (38.93, 175.85) | 90.41 (42.56, 168.25) | 79.21 (31.30, 187.18) | 0.329 |
| **C-reactive protein (mg/L)** | 146.60 (71.80, 226.10) | 147.90 (78.03, 230.40) | 142.00 (67.50, 216.05) | 0.203 |
| **Alkaline phosphatase (U/L)** | 81.00 (60.00, 111.25) | 80.50 (60.00, 104.75) | 81.00 (61.00, 118.50) | 0.063 |
| **Glutamyl transpeptidase (U/L)** | 55.00 (28.00, 123.00) | 51.00 (28.50, 106.50) | 57.50 (27.25, 144.00) | 0.013 |
| **Total protein (g/L)** | 57.00 (51.80, 63.40) | 57.10 (51.65, 63.32) | 56.90 (52.35, 63.35) | 0.966 |
| **Glutamate dehydrogenase (U/L)** | 4.00 (2.00, 10.00) | 3.00 (2.00, 7.75) | 5.00 (3.00, 13.00) | 0.093 |
| **Alanine aminotransferase (U/L)** | 27.00 (18.00, 43.00) | 27.00 (18.00, 42.00) | 28.00 (18.75, 44.25) | 0.358 |
| **Indirect bilirubin (μmol/L)** | 7.85 (4.40, 13.12) | 8.10 (4.43, 12.97) | 7.65 (4.40, 13.70) | 0.922 |
| **Albumin (g/L)** | 30.80 (27.70, 34.20) | 30.45 (27.55, 34.70) | 30.80 (27.85, 33.70) | 0.713 |
| **Total bilirubin (μmol/L)** | 17.60 (10.90, 29.70) | 16.85 (11.35, 28.12) | 18.10 (10.75, 32.10) | 0.148 |
| **Direct bilirubin (μmol/L)** | 0.20 (0.00, 6.30) | 0.10 (0.00, 5.55) | 0.40 (0.00, 6.50) | 0.135 |
| **Total bile acids (μmol/L)** | 4.10 (3.40, 5.70) | 4.10 (3.48, 5.55) | 4.00 (3.40, 5.73) | 0.975 |
| **Aspartate aminotransferase (U/L)** | 28.00 (19.00, 49.00) | 27.50 (19.00, 51.00) | 30.00 (19.00, 48.00) | 0.719 |
| **Retinol binding protein (mg/L)** | 18.00 (11.00, 30.00) | 18.00 (12.00, 29.75) | 18.50 (10.00, 32.25) | 0.278 |
| **Lactate dehydrogenase (U/L)** | 850.00 (542.00, 1433.50) | 896.50 (555.25, 1539.25) | 806.00 (528.00, 1362.00) | 0.877 |
| **Glucose (mmol/L)** | 8.10 (6.00, 11.90) | 8.10 (6.10, 11.75) | 7.95 (5.97, 11.93) | 0.613 |
| **Prealbumin (mg/L)** | 101.00 (79.00, 126.00) | 99.00 (76.25, 120.75) | 106.00 (80.25, 133.75) | 0.116 |
| **Globulin (g/L)** | 26.30 (22.70, 29.90) | 26.00 (22.83, 29.82) | 26.50 (22.70, 29.80) | 0.66 |
| **Blood urea nitrogen (mmol/L)** | 4.75 (3.00, 9.65) | 4.70 (3.00, 7.80) | 5.40 (3.30, 11.60) | 0.004 |
| **Creatinine (μmol/L)** | 53.31 (39.58, 103.95) | 52.00 (39.25, 73.00) | 59.00 (40.90, 146.15) | 0.063 |
| **Cystatin C (mg/L)** | 0.68 (0.52, 1.17) | 0.60 (0.49, 0.86) | 0.80 (0.61, 1.81) | <0.001 |
| **Brain natriuretic peptide precursor (pmol/L)** | 23.88 (7.39, 76.83) | 20.48 (6.00, 61.77) | 30.25 (10.10, 108.00) | 0.282 |
| **Creatine kinase MB (U/L)** | 3.00 (3.00, 6.00) | 3.00 (3.00, 7.00) | 3.00 (3.00, 5.00) | 0.797 |
| **Creatine kinase (U/L)** | 50.00 (20.00, 165.50) | 52.00 (20.00, 233.00) | 42.50 (20.00, 148.75) | 0.525 |
| **Myohemoglobin (ng/ml)** | 44.00 (20.95, 201.40) | 42.95 (19.92, 181.45) | 48.90 (21.15, 223.35) | 0.312 |
| **TroponinT (ng/ml)** | 0.01 (0.01, 0.03) | 0.01 (0.00, 0.03) | 0.01 (0.01, 0.03) | 0.369 |
| **TroponinI (ng/ml)** | 0.03 (0.02, 0.05) | 0.03 (0.02, 0.04) | 0.03 (0.02, 0.05) | 0.182 |
| **Sodium (mmol/L)** | 137.90 (134.83, 140.00) | 137.50 (134.10, 140.00) | 138.00 (135.30, 140.00) | 0.035 |
| **Magnesium (mmol/L)** | 0.78 (0.66, 0.87) | 0.79 (0.67, 0.85) | 0.75 (0.65, 0.88) | 0.979 |
| **Chlorine (mmol/L)** | 102.00 (98.00, 106.00) | 102.00 (98.00, 106.00) | 102.00 (98.00, 105.00) | 0.836 |
| **Phosphorus (mmol/L)** | 1.00 (0.77, 1.29) | 0.98 (0.70, 1.19) | 1.12 (0.81, 1.35) | 0.001 |
| **Potassium (mmol/L)** | 4.00 (3.70, 4.40) | 4.00 (3.70, 4.30) | 4.10 (3.80, 4.40) | 0.081 |
| **Calcium (mmol/L)** | 2.07 (1.88, 2.20) | 2.03 (1.87, 2.18) | 2.11 (1.90, 2.26) | 0.031 |
| **Apolipoprotein E (mg/L)** | 79.45 (61.88, 100.83) | 77.10 (64.62, 102.25) | 81.50 (54.75, 95.50) | 0.496 |
| **Apolipoprotein B (g/L)** | 0.79 (0.52, 1.11) | 0.80 (0.60, 1.12) | 0.79 (0.48, 1.10) | 0.49 |
| **Apolipoprotein A (g/L)** | 0.66 (0.51, 0.83) | 0.67 (0.55, 0.79) | 0.64 (0.46, 0.89) | 0.192 |
| **Free fatty acid (mmol/L)** | 0.51 (0.37, 0.64) | 0.51 (0.31, 0.64) | 0.51 (0.38, 0.69) | 0.711 |
| **HDL cholesterol (mmol/L)** | 0.45 (0.32, 0.66) | 0.47 (0.33, 0.64) | 0.43 (0.30, 0.69) | 0.901 |
| **Total cholesterol (mmol/L)** | 3.70 (2.62, 5.80) | 3.82 (2.82, 6.19) | 3.42 (2.34, 5.53) | 0.056 |
| **Triglycerides (mmol/L)** | 3.86 (2.41, 5.90) | 3.82 (2.50, 5.90) | 3.90 (2.24, 5.90) | 0.18 |
| **LDL cholesterol (mmol/L)** | 1.98 (1.16, 3.32) | 2.08 (1.27, 3.53) | 1.85 (1.13, 3.09) | 0.691 |
| **Lipase (U/L)** | 360.00 (116.00, 742.50) | 411.00 (146.00, 971.50) | 327.00 (105.00, 601.75) | 0.061 |
| **Amylase (U/L)** | 79.00 (41.00, 163.75) | 84.50 (40.75, 191.00) | 72.50 (41.00, 127.00) | 0.227 |
| **Insulin (mIU/L)** | 5.76 (3.45, 14.13) | 7.17 (5.18, 8.75) | 4.64 (3.25, 16.57) | 0.51 |
| **Glycated hemoglobin** | 6.00 (5.40, 7.57) | 6.05 (5.50, 7.70) | 5.90 (5.27, 7.15) | 0.362 |
| **C-Peptide (ng/L)** | 1.69 (0.71, 3.70) | 1.69 (0.84, 3.77) | 1.73 (0.45, 3.11) | 0.829 |
| **Free thyroid hormone 3 (pmol/L)** | 2.87 (2.44, 3.24) | 2.88 (2.48, 3.23) | 2.87 (2.39, 3.24) | 0.625 |
| **Free thyroid hormone 4 (pmol/L)** | 9.80 (8.50, 11.77) | 9.70 (8.50, 11.17) | 10.00 (8.20, 12.10) | 0.244 |
| **Thyroid hormone 3 (nmol/L)** | 0.49 (0.35, 0.69) | 0.48 (0.32, 0.65) | 0.51 (0.40, 0.72) | 0.097 |
| **Calcitonin (pg/ml)** | 6.06 (4.06, 6.89) | 6.20 (4.09, 6.98) | 5.51 (4.05, 6.85) | 0.57 |
| **Thyroid hormone 4 (nmol/L)** | 67.69 (51.84, 87.20) | 68.62 (53.17, 82.50) | 67.05 (51.38, 94.29) | 0.58 |
| **Thyroglobulin (ug/L)** | 3.90 (1.88, 7.45) | 3.72 (2.21, 7.07) | 4.18 (0.95, 8.30) | 0.328 |
| **Parathyroid Hormone (pg/ml)** | 4.30 (2.20, 8.50) | 4.25 (2.10, 8.65) | 4.40 (2.40, 8.10) | 0.646 |
| **Carbohydrate antigenCA125 (U/ml)** | 37.45 (18.40, 94.43) | 37.60 (18.40, 88.43) | 34.92 (23.86, 101.50) | 0.69 |
| **Carbohydrate antigenCA199 (U/ml)** | 13.08 (8.68, 26.81) | 12.25 (8.68, 21.26) | 18.18 (9.12, 28.63) | 0.352 |

Descriptive statistics are presented as median (IQR) for continuous laboratory measures, and frequency (percentage, %) for categorical laboratory measures.

*p*-values were derived from Mann-Whitney U test for continuous laboratory measures.

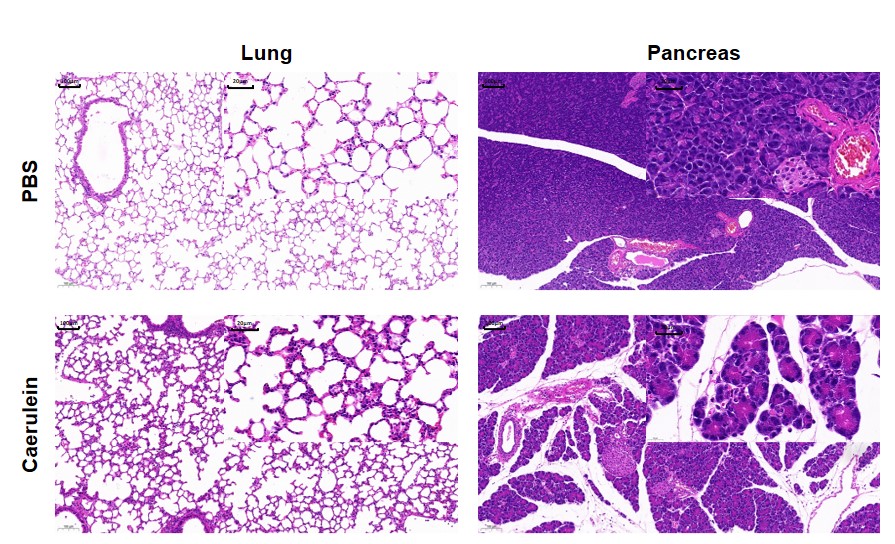
**Supplementary Fig. 1 Pathology examination of lung and pancreas tissue in mice.**

The pathological examination of mice induced as acute pancreatitis presented acinar cell necrosis, endo-cellular and extra-cellular edema, infiltration of neutrophils in pancreas tissue, and severe damage of alveoli. Abbreviations: PBS, Phosphate Buffer Saline.

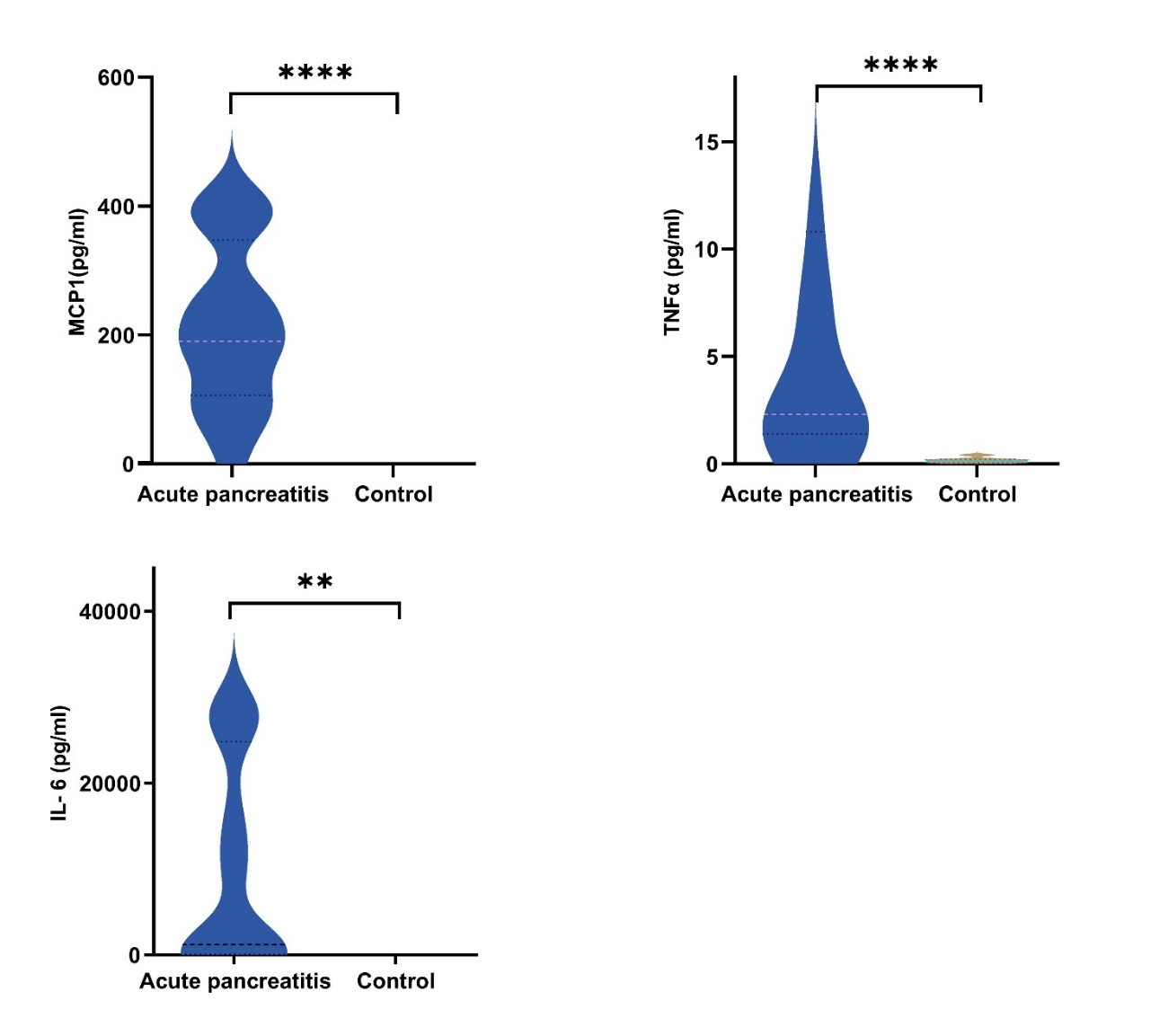

**Supplementary Fig. 2 Assays of blood MCP1, TNFα and IL-6 from mice induced as AP and normal healthy mice (n=10 in each group).** ** indicates p<0.01, **** indicates p<0.0001 by Mann-Whitney test.

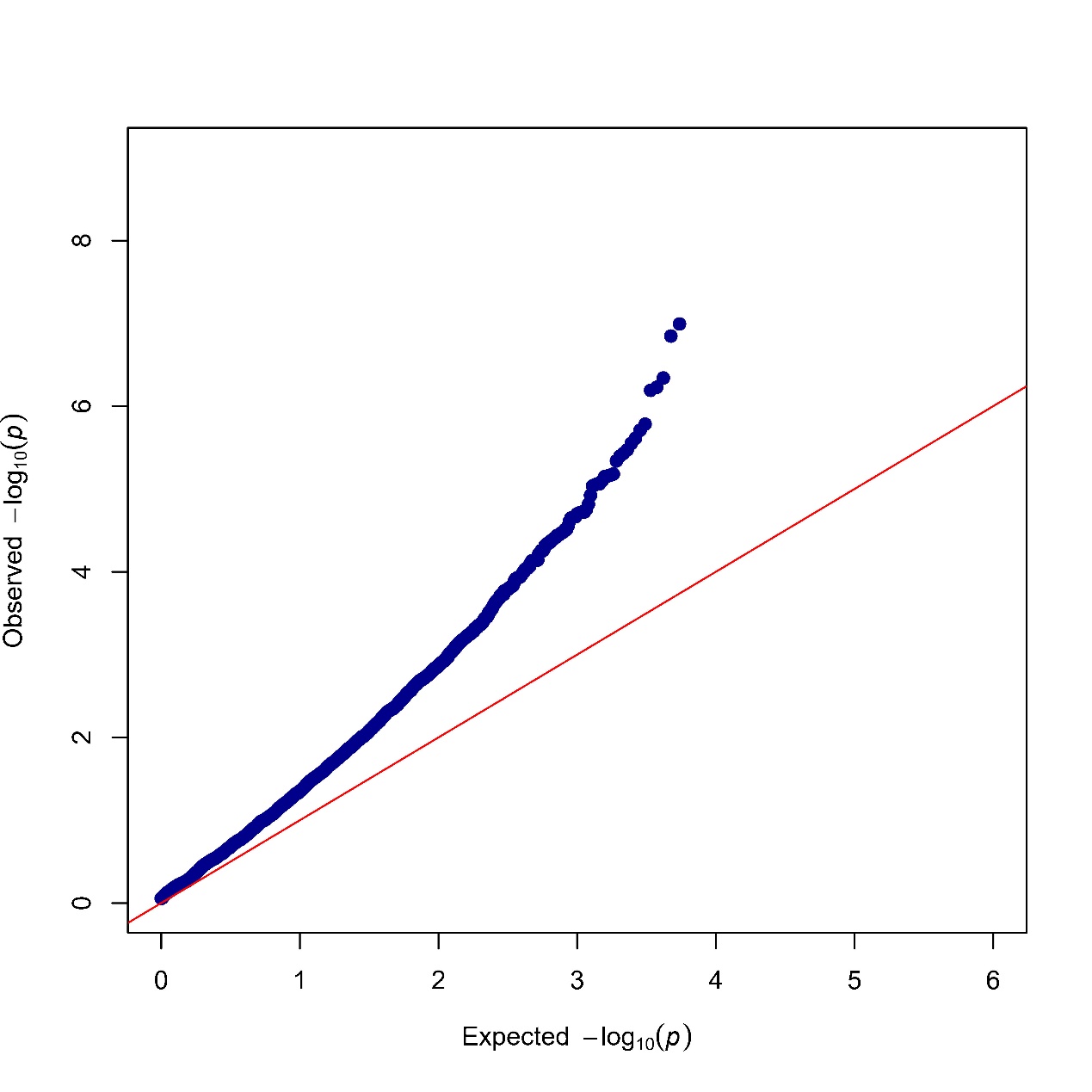

**Supplementary Fig. 3 Quantile-quantile plot for the GWAS analysis of WES and SIH.**

The y axis indicates the actual -log_10_ (*p* value of each variant) and the x axis indicates the expected -log10 (*p* value of each variant).

Abbreviations: GWAS, genome-wide association study; WES, whole exome sequence; SIH, spontaneous intraabdominal hemorrhage.

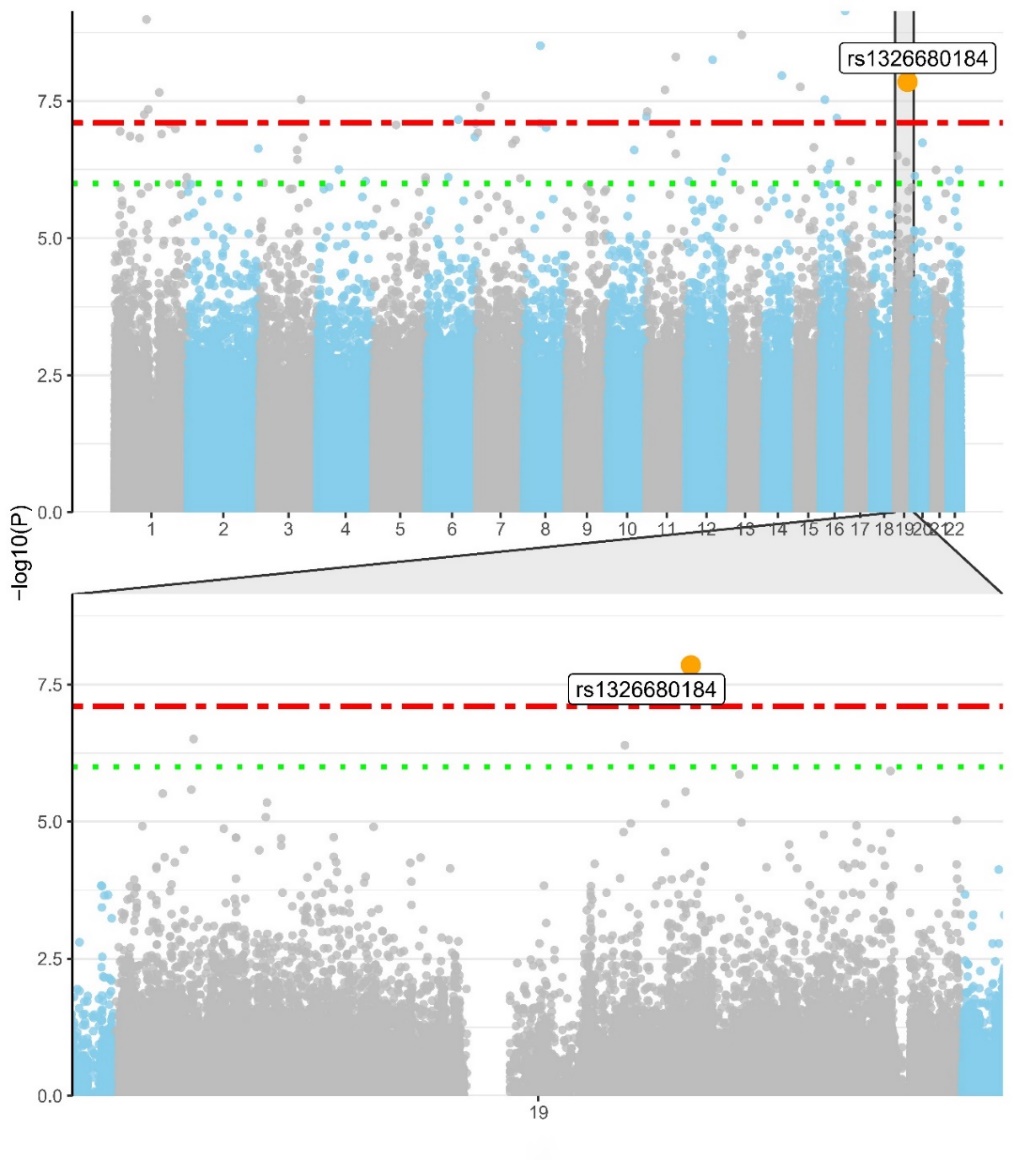

**Supplementary Fig. 4 Manhattan plot of genome-wide association analysis for exonic variants in all autosomal chromosomes.**

The genome-wide significance was set at 1×10^-6^ (1×10^-8^/1×10^-2^ as exome accounts for approximately 1% of human genome).

The rs1326680184 was used and denoted to represent the identified six FCGBP variants.

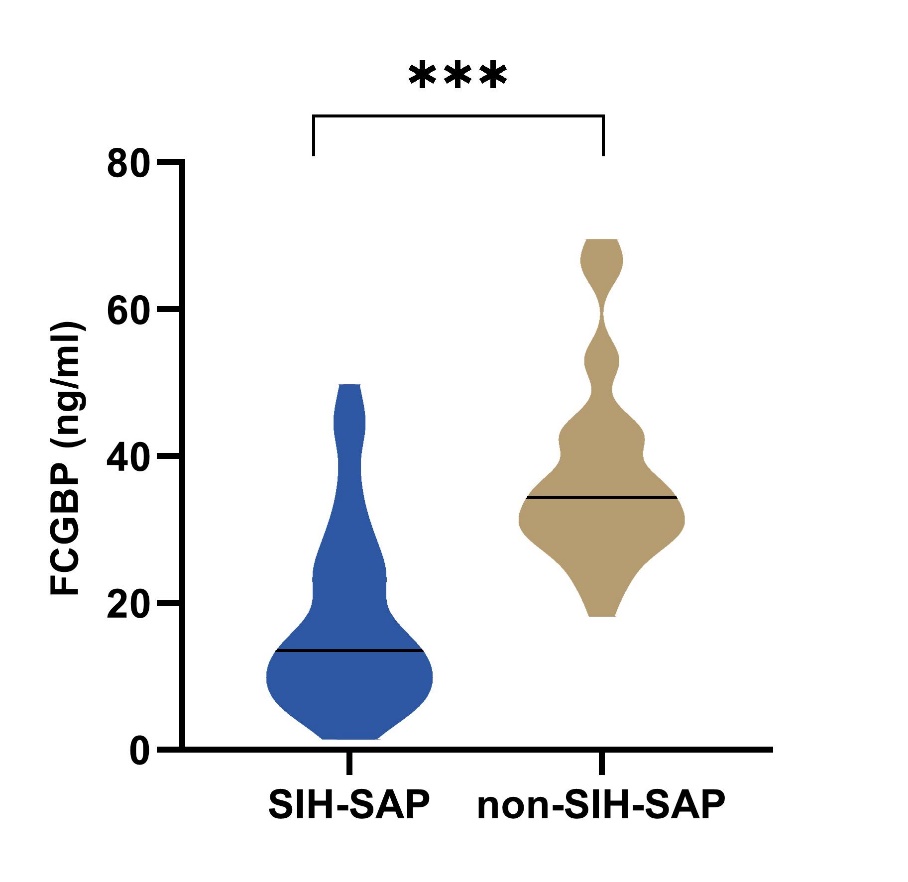

**Supplementary Fig. 5 FCGBP protein level of patients with or without SIH.**

*** indicates p<0.001 by Mann-Whitney test.

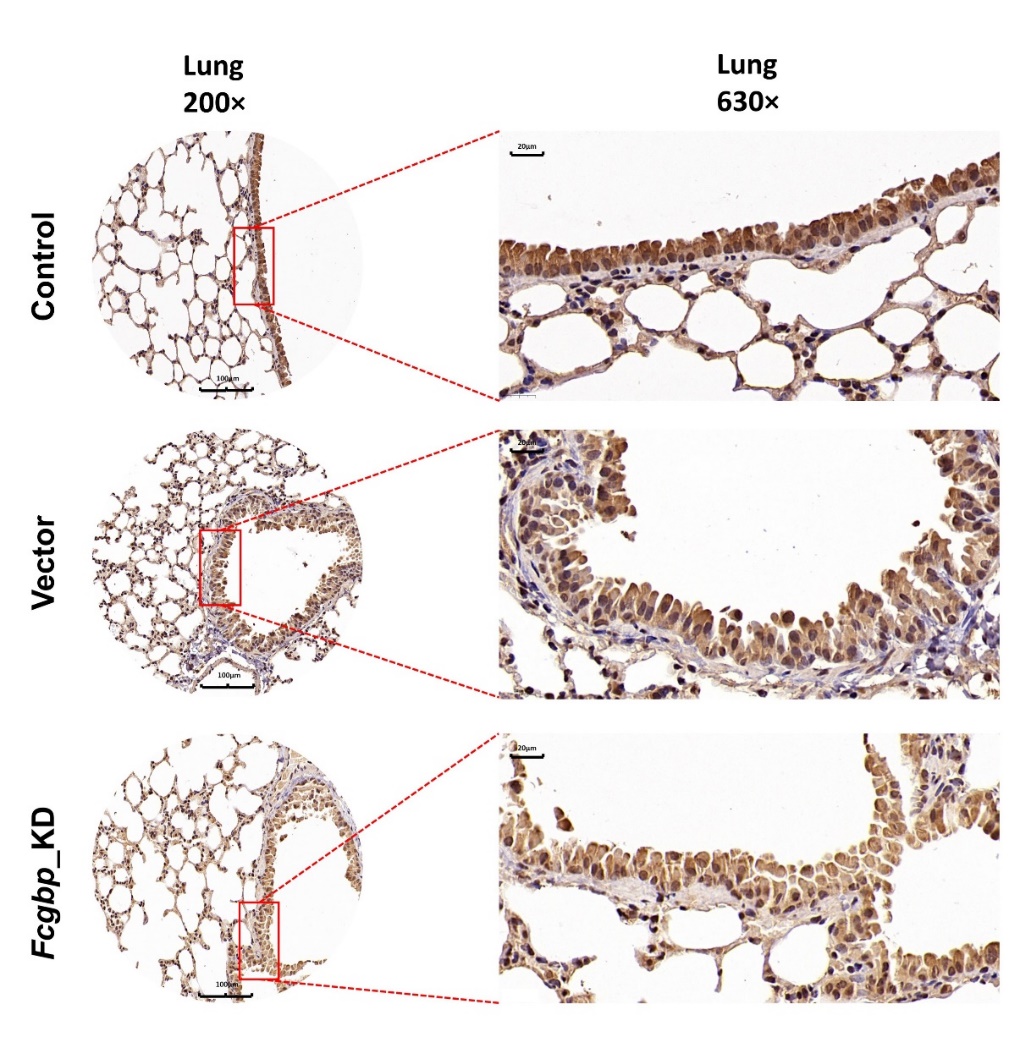

**Supplementary Fig. 6 Immunohistochemical analysis of expression of *Fcgbp* in lung tissue in mice.**

Representative IHC images of Fcgbp in lung tissue were shown (n=10 in each group), which indicates the injection of AAV-RNAi successfully reduces the expression of *Fcgbp* levels in lung tissue in mice.

Abbreviations: IHC, Immunohistochemistry; AAV-RNAi, adeno-associated virus harboring short.

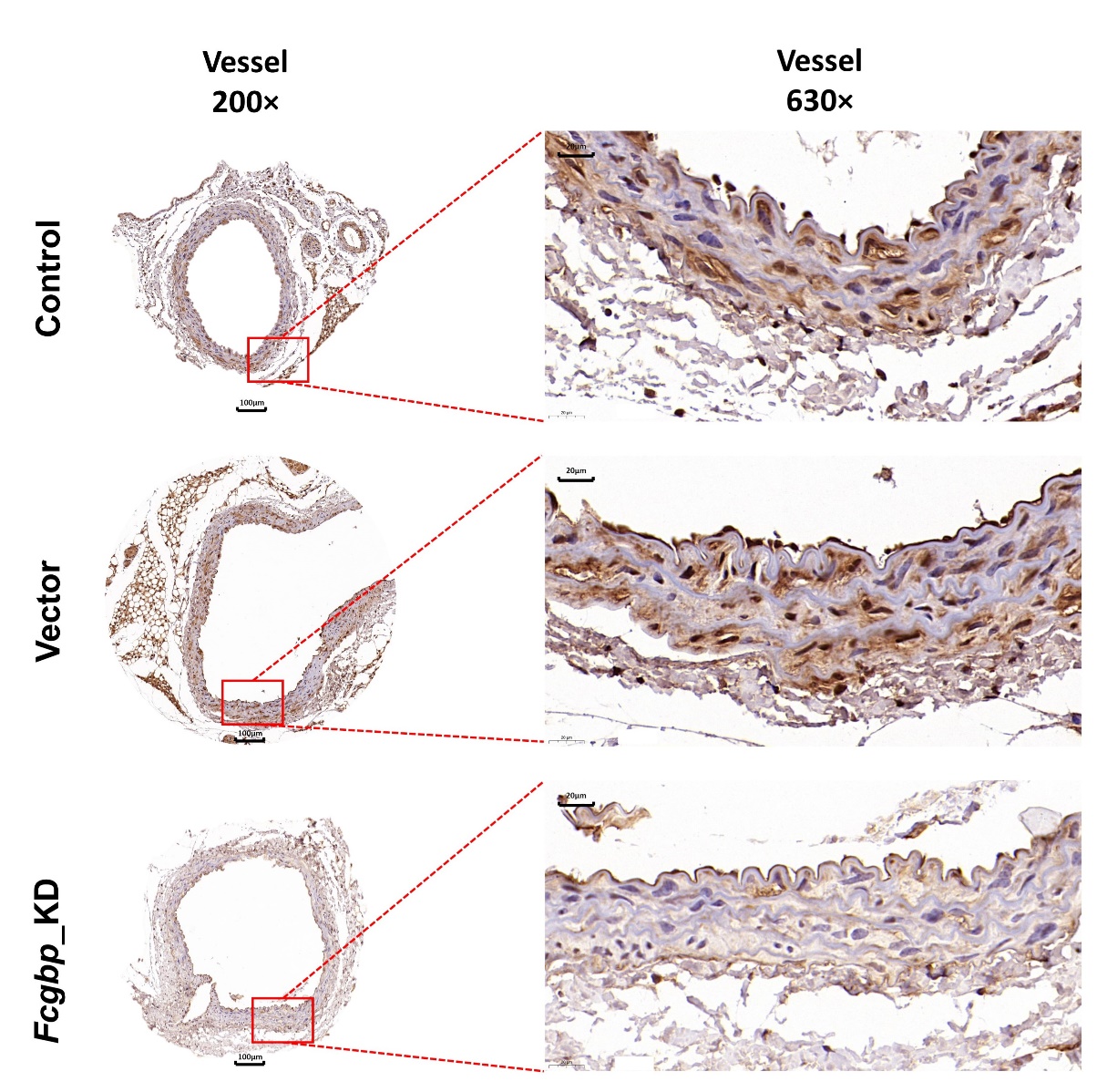

**Supplementary Fig. 7 Immunohistochemical analysis of *Fcgbp* expression in vessel tissue in mice.**

Representative IHC images of Fcgbp in vessel tissue were shown (n=10 in each group), which indicates the injection of AAV-RNAi successfully reduces the expression of *Fcgbp* levels in tissue of vessel in mice.

Abbreviations: IHC, Immunohistochemistry; AAV-RNAi, adeno-associated virus harboring short.

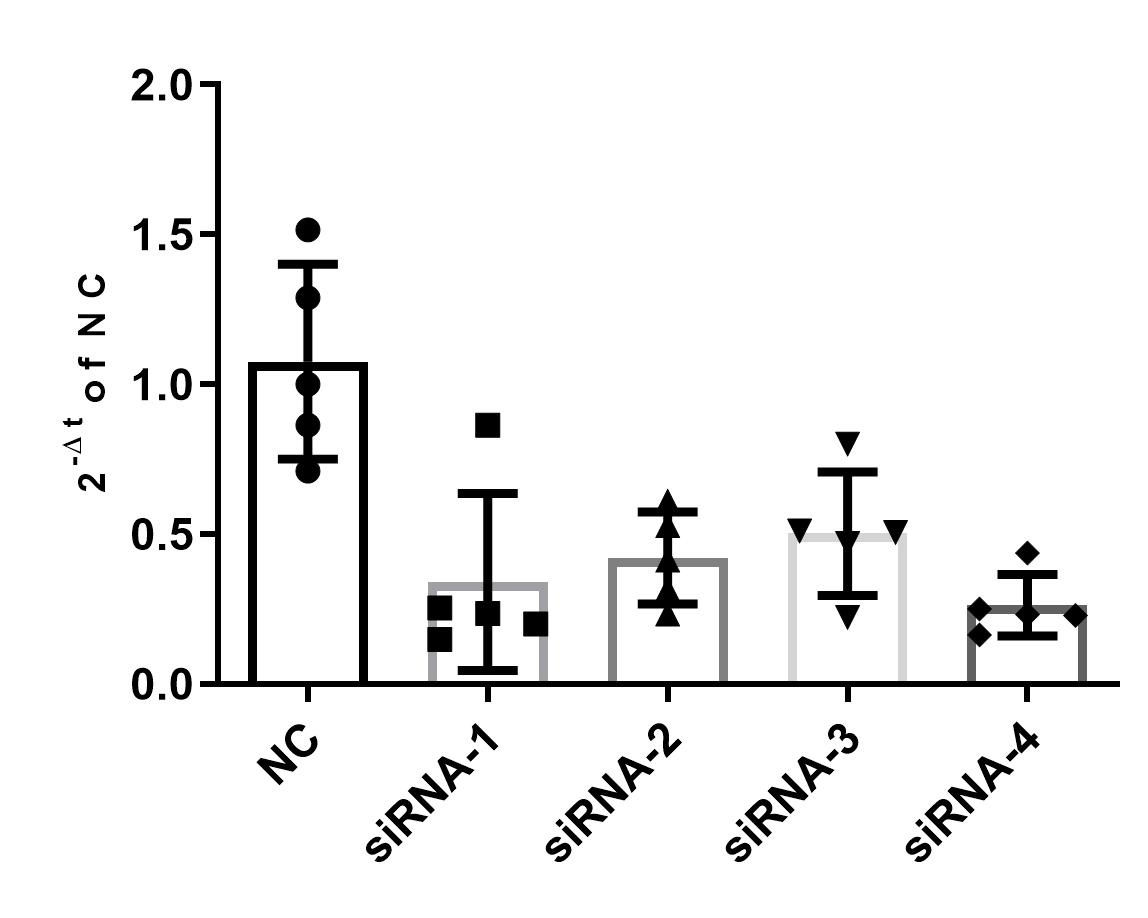

**Supplementary Fig. 8 Quantitative PCR analysis of FCGBP mRNA level in knockdown fibroblasts and wild-type control.**
